# A Flexible Framework for Local-Level Estimation of the Effective Reproductive Number in Geographic Regions with Sparse Data

**DOI:** 10.1101/2024.11.06.24316859

**Authors:** Md Sakhawat Hossain, Ravi Goyal, Natasha K Martin, Victor DeGruttola, Mohammad Mihrab Chowdhury, Christopher McMahan, Lior Rennert

## Abstract

**Background:** Our research focuses on local-level estimation of the effective reproductive number, which describes the transmissibility of an infectious disease and represents the average number of individuals one infectious person infects at a given time. The ability to accurately estimate the infectious disease reproductive number in geographically granular regions is critical for disaster planning and resource allocation. However, not all regions have sufficient infectious disease outcome data; this lack of data presents a significant challenge for accurate estimation.

**Methods:** To overcome this challenge, we propose a two-step approach that incorporates existing *R*_*t*_ estimation procedures (EpiEstim, EpiFilter, EpiNow2) using data from geographic regions with sufficient data (step 1), into a covariate-adjusted Bayesian Integrated Nested Laplace Approximation (INLA) spatial model to predict *R*_*t*_ in regions with sparse or missing data (step 2). Our flexible framework effectively allows us to implement any existing estimation procedure for *R*_*t*_ in regions with coarse or entirely missing data. We perform external validation and a simulation study to evaluate the proposed method and assess its predictive performance.

**Results:** We applied our method to estimate *R*_*t*_ using data from South Carolina (SC) counties and ZIP codes during the first COVID-19 wave (‘Wave 1’, June 16, 2020 – August 31, 2020) and the second wave (‘Wave 2’, December 16, 2020 – March 02, 2021). Among the three methods used in the first step, EpiNow2 yielded the highest accuracy of *R*_*t*_ prediction in the regions with entirely missing data. Median county-level percentage agreement (PA) was 90.9% (Interquartile Range, IQR: 89.9-92.0%) and 92.5% (IQR: 91.6-93.4%) for Wave 1 and 2, respectively. Median zip code-level PA was 95.2% (IQR: 94.4-95.7%) and 96.5% (IQR: 95.8-97.1%) for Wave 1 and 2, respectively. Using EpiEstim, EpiFilter, and an ensemble-based approach yielded median PA ranging from 81.9%-90.0%, 87.2%-92.1%, and 88.4%-90.9%, respectively, across both waves and geographic granularities.

**Conclusion:** These findings demonstrate that the proposed methodology is a useful tool for small-area estimation of *R*_*t*_, as our flexible framework yields high prediction accuracy for regions with coarse or missing data.

## Introduction

Infectious disease outbreaks pose significant threats, impacting health, operations, and the economy. Effective outbreak management relies on the ability to estimate the intensity of local transmission and strategize appropriately, which is vital for controlling disease transmission, enhancing health outcomes, and reducing economic impacts. However, current approaches often target broad geographic areas, which may not yield actionable insights for local-level or institutional decision-making [1]. This lack of granularity can be attributed to variations in community characteristics such as age distribution, prevalence of comorbidities, and social behaviors [2,3], all of which influence disease transmission and susceptibility to severe outcomes. Moreover, policies derived from coarse data may lead to suboptimal local disease control [4,5]. Therefore, enhancing local-level transmission estimation and forecasting is essential for tailoring policies and optimizing interventions.

The effective reproductive number is a key epidemiological metric used to describe the transmissibility of infectious diseases for a specific time and setting; it is critical to understand the intensity of transmission. *R*_*t*_ represents the average number of people infected by one infectious individual at a given time *t*. An *R*_*t*_ value greater than 1 signifies that the disease is spreading, while an *R*_*t*_ less than 1 indicates that the transmission is declining [6,7]. *R*_*t*_ changes with the progression of an epidemic and the implementation of control measures [8–10]. It also varies spatially due to the differences in population density, mobility, social interactions, and public health policies [11,12]. Unfortunately, local-level estimation of *R*_*t*_ is challenging due to data sparsity or unavailability of infectious disease testing or encounter records. For example, insufficient testing and changes in case definitions or reporting can substantially impact estimates of *R*_*t*_ [13,14]. Due to substantial lags in producing estimates of disease epidemiological metrics from traditional surveillance systems [15], electronic health records (EHR) obtained directly from health systems or hospitals are useful for real-time estimation of such metrics, including *R*_*t*_. However, healthcare systems do not serve all populations, which inhibits the ability to forecast disease spread in communities that are not represented in the healthcare system. Additionally, estimates of *R*_*t*_ may be unstable for communities that are underrepresented in the healthcare system and hence offer few data points for accurate estimation.

There are several techniques for estimating the effective reproductive number; e.g., Wallinga and Tenius method [16], Cori et al. method [17], EpiFilter [6], and EpiNow2 [18]. EpiEstim is a tool integrated into widely used software platforms for estimation of the reproductive number from incidence time series, accounting for uncertainties in the serial interval distribution [17]. EpiFilter utilizes all available incidence information and does not rely on window size assumptions, making it more statistically robust, particularly during periods of low incidence [6]. The modeling approach of EpiNow2, an R software package [18], is based on the methods developed by Cori et al. and Thompson et al. [19], and it incorporates the delays from infection to reporting to estimate the effective reproductive number utilizing a range of open-source tools [20]. Although there are several methods for estimating the effective reproductive number, there is a notable gap in accurately estimating *R*_*t*_ for small areas with coarse or missing data. To address this gap, we develop a flexible framework that effectively allows us to implement any estimation procedure for *R*_*t*_ (e.g., EpiEstim, EpiFilter, EpiNow2, etc.) in regions with coarse or entirely missing data.

The motivation for this study, funded by the National Library of Medicine of the National Institutes of Health (NIH) and the Center for Forecasting and Outbreak analytics of the Centers for Disease Control and Prevention (CDC), is to inform allocation of mobile health clinics for infectious disease interventions to medically underserved communities at high risk of infectious disease outbreaks. We address the challenge of local level *R*_*t*_ estimation in areas with coarse or entirely missing data by proposing a two-step estimation procedure which incorporates existing *R*_*t*_ estimation techniques along with an ensemble estimation using data from geographic regions with sufficient data (step 1) into a spatial modeling framework to predict *R*_*t*_ in regions with sparse or missing data (step 2).

The remainder of the paper is organized as follows: the **Materials and Methods** section outlines the framework of the study, detailing the statistical techniques and data utilized. In the **Results** section, we present our findings and interpret the results. The **Discussion** section explores the implications of the results, acknowledges the limitations of our study, and suggests directions for future research. The paper concludes with the **Conclusion** section, summarizing the main points.

## Materials and Methods

To estimate the region-specific effective reproductive number (*R*_*t*_) for an infectious disease in small areas with coarse or entirely missing data, we develop a two-step procedure. In the first step of our analyses, we estimate *R*_*t,i*_, using the existing techniques EpiEstim, EpiFilter, and EpiNow2 for each geographic region *i* [6,17,18]. We denote the resulting estimates by 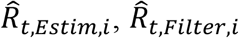, and 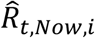, respectively. We also perform an ensemble-based estimation 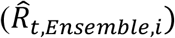 by considering equal weights for each estimation technique. In the second step, we employ a Bayesian Integrated Laplace Approximation (INLA) model incorporating spatial and sociodemographic information. For each geographic region *i* with available outcome data (e.g., Covid-19 cases), we denote these estimates by 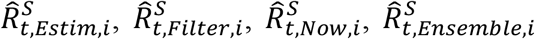, which are referred to as spatially and covariates smoothed estimates. When geographic region *i* is not included in the spatial (and covariate-adjusted) INLA model fitting in step 2 (e.g., geographic region *i* has entirely missing data), we denote the prediction in this region by 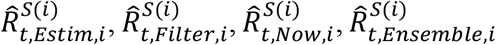, where *S*(*i*) indicates information from region *i* is not used in the model fitting in step 2. More details of initial estimation, smoothing and predictions of *R*_*t*_ in regions with entirely missing data, along with their evaluations, are discussed in the respective sections.

### Data Description

The number of cumulative confirmed COVID-19 case data for US counties is publicly available on the New York Times (NYT) GitHub repository [21]. The daily reported cases are calculated from the county-level cumulative data. We obtained the ZIP code level daily COVID-19 case data from Prisma Health’s (SC’s largest not-for-profit health care provider) COVID-19 registry. We utilize data from SC counties and ZIP codes during the first COVID-19 wave (Wave 1, between June 16, 2020 – August 31, 2020) and the second wave (Wave 2, between December 16, 2020 – March 02, 2021). Since many ZIP codes in the Prisma Health system data lack sufficient case numbers, we restrict our analysis to those ZIP codes with at least 50 cases in each period to ensure the reliability of our results (Wave 1: N = 30 ZIP codes; Wave 2: N = 45 ZIP codes). Population demographics are characterized by age, sex, race/ethnicity, employment, and insurance coverage. These data are sourced from the United States Census Bureau website [22]. The social vulnerability index (SVI) data is obtained from the Agency for Toxic Substances and Disease Registry website [23].

### Initial Estimation of *R*_*t*_

We utilize the techniques EpiEstim, EpiFilter, and EpiNow2 to obtain the initial estimates of the effective reproductive number. EpiEstim and EpiFilter assume the incidence *I*_*t*_ at time *t* is Poisson distributed with mean 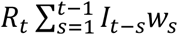, where *R*_*t*_ is the effective reproductive number at time *t, s* is the time since infection of the case, *w*_*s*_ is the probability that a primary case takes time between *s* − 1 and *s* days to generate the secondary infection, and *w*_*s*_ is obtained from the serial interval distribution [6,17]. EpiEstim uses an expectation-maximization algorithm to reconstruct daily incidence from aggregated data for estimating *R*_*t*_ [24]. It computes estimates of the effective reproductive number, 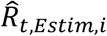, using a sliding time window; the default seven days window setting is used. EpiFilter employs a recursive Bayesian smoothing technique to estimate the effective reproductive number, 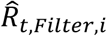. It unifies and extends two popular techniques EpiEstim and the Wallinga-Teunis method [6]. EpiNow2 employs a Bayesian latent variable model that incorporates a probabilistic programming language, Stan [20,25]. Detailed description of the EpiEstim, EpiFilter, and EpiNow2 methods are available in the literature [6,17,18,20]. Each of the three methods estimates *R*_*t*_ using different parameters or data assumptions. All three methods depend on the generation time distribution. Since infection times are often unobserved, the generation time is usually approximated using the serial interval distribution [17]. In all three methods, the parameters for both the serial interval distribution and incubation period are utilized.

From the literature, we obtain a mean of 5 days (95% CI: 4.94-5.06) and a standard deviation (SD) of 2.4 for the incubation period [26], a mean of 4.7 days and an SD of 2.9 for the serial interval distribution [27]. The ‘estimate_R’ function of the EpiEstim, R software package version 2.2.4 is used to obtain 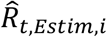. We calculate the estimate, 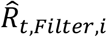, by using the R software function ‘epiSmoother’, available at https://github.com/kpzoo/EpiFilter. The ‘epinow’ function from the R software package EpiNow2, version 1.4.0, is utilized to get 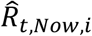. We develop an ensemble method to obtain 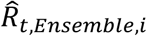 by averaging 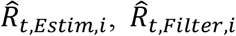, and 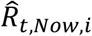. The ensemble method aims to reduce the bias and variance in epidemiological forecasting by averaging the various model outputs [28].

### Proposed Covariate-Adjusted Spatial Smoothing via INLA

We propose a sociodemographic covariate adjusted, two-step Integrated Nested Laplace Approximation (INLA) model to refine the initial estimation of the effective reproductive number at the local-level and to provide estimates for the regions with coarse or missing data. The use of INLA for spatial data prediction is supported by extensive research, demonstrating its effectiveness in handling spatial dependency and missing data challenges[29,30]. The INLA method is designed to derive the posterior marginal distribution for latent Gaussian models and serves as a computationally efficient alternative to traditional Markov Chain Monte Carlo (MCMC) methods for Bayesian inference [29,31]. INLA utilizes a series of nested Laplace approximations to calculate the marginal posterior and hyperparameters of latent Gaussian models. The INLA framework is employed to integrate the spatial dependencies of the effective reproductive number estimates into our model [32]. To fit the INLA model, we use the R software package called “R-INLA” [29,30,33]. We incorporate the effect of the spatial location (counties or ZIP codes) in the INLA function through Besag’s model [34,35]. The Besag model employs spatial random effects for each area to capture the correlation among neighboring areas, adjusting the estimated *R*_*t*_ based on the estimates of the neighboring areas. Neighborhood structures are defined using the ‘poly2nb()’ function, identifying areas as neighbors if their boundary polygons share at least one vertex. We then convert the neighborhood structure into a binary adjacency matrix using the ‘nb2mat()’ function. Each element of the adjacency matrix represents whether two locations are neighbors (1 if they share a boundary, 0 if they do not).

The model equation in INLA is:

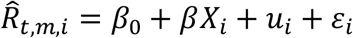

where *β*_0_ is the intercept of the model, *β* is the coefficients vector for sociodemographic community-level covariates 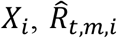 corresponds to the estimates 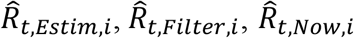, or 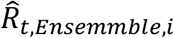 at area *i*.The term *u*_*i*_ specifies the spatial random effect for location *i*, which depends on the neighboring locations defined by the adjacency matrix, and the random Gaussian error term is *ε*_*i*_.

The sociodemographic covariates used for the analyses at the county and ZIP code levels are the percentage of the population for different age groups (“Age 0-19”, “Age 20-44”, “Age 45-64”, “Age 65 and over”), the percentage of the population for different races and ethnicity (“White”, “Hispanic”, “Black or African American” and “others”), the percentage of the population “Employed” and “Unemployed”, the percentage of “Health Insurance Coverage”, the percentage of the population of “Male” and “Female”, the Social Vulnerability Index (SVI), and household median income. Household median income is normalized from 0 to 1, and SVI values range from 0 to 1, where higher values indicate greater social vulnerability. We consider the covariates “Age 0-19”, “White”, “Unemployed”, “Uninsured”, and “Female” population groups as the reference categories.

### Assessment of Prediction of *R*_*t*_ for Regions with Entirely Missing Data

We employ the two-step spatial (covariate-adjusted) INLA model using the initial estimate, 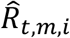, for each method *m* (i.e., EpiEstim, EpiFilter, EpiNow2, and Ensemble) at location *i*, as the response variable. We validate the model using two approaches. In the first approach, we randomly select 90% of the regions to hold out for the training set and 10% for the test set. We then apply the proposed two-step spatial (covariate-adjusted) INLA model to the training set and use this model to predict the effective reproductive number for the geographic regions in the test set, which we denote by 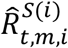. We then compare the agreement between 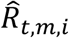, and 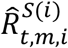 for the geographic regions that are left out from the model fitting in the second step. The prediction accuracy measurement metrics are calculated using the formulas in equations 1-3. This process is repeated 200 times.

In the second validation approach, we hold out geographic regions with sufficient data for the test set – specifically, Charleston, Greenville, Horry, and Richland counties and ZIP codes 29605, 29642, 29680, and 29681. Since there is no gold standard for comparing *R*_*t*_ estimates, we select the above regions to predict *R*_*t*_ for test cases with entirely missing data, assuming the initial estimates from regions with sufficient data will serve as a baseline for comparison. The prediction in this approach is performed by leaving out one area at a time.

The prediction accuracy measurement metrics are calculated using the following formulas:

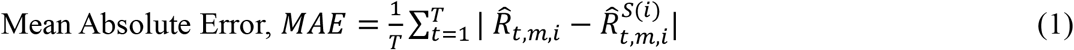

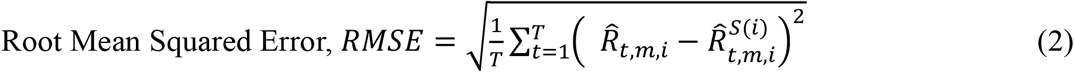

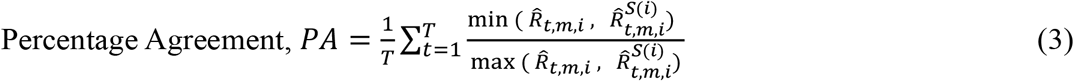

where,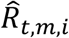 is the initial estimates, 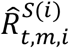 is the prediction of effective reproductive number for each method *m*, at time *t* and location *i*, and *T* is the total number of time points (days).

### Simulation Study

We conduct a simulation study to validate our two-step spatial (covariate-adjusted) INLA framework under controlled conditions. To generate realistic effective reproductive number (*R*_*t*_) values, we capture the time-varying nature of disease transmission using a sinusoidal function. The parameters for the sinusoidal function are drawn from a multivariate normal (MVN) distribution, with a covariance structure derived from the adjacency matrix of the counties in SC. This structure ensures that spatial correlations are incorporated, allowing neighboring counties to have similar transmission patterns. The parameters are then scaled to fit epidemiologically meaningful ranges, representing the baseline reproductive number, amplitude, frequency of disease waves, and linear adjustments. Using these parameters, *R*_*t*_ values are generated and used to simulate daily COVID-19 case counts at the county level. The number of new infections at time *t* is based on total infectiousness, which depends on past cases weighted by a discretized gamma-distributed serial interval.

To evaluate the performance of different estimation methods, we generate 50 independent sets of daily COVID-19 case data. The analysis follows the same workflow as for real data, including initial *R*_*t*_ estimation, two-step spatial (covariate-adjusted) INLA smoothing, and prediction of *R*_*t*_ for counties with entirely missing data. For initial estimation, we apply EpiEstim, EpiFilter, and an ensemble-based approach, excluding EpiNow2 due to its high computational cost. To assess the predictive performance of our framework, we systematically remove selected counties from the INLA model fitting process, making their data unavailable. The model is then trained using data from the remaining counties, and *R*_*t*_ is predicted for the excluded counties. Unlike real data, where the true *R*_*t*_ is unknown, the simulation study generates true *R*_*t*_ values, allowing to benchmark and compare the performance of different estimation methods. Further details on the simulation methodology are provided in the **Supporting Information 1 (pages 14–17)**.

## Results

### Initial Estimation and Spatial (covariate-adjusted) INLA Smoothing of *R*_***t***_

For the *R*_*t*_ estimation in this section, we utilized the county and ZIP code level COVID-19 daily case data for the first wave (June 16, 2020, through August 31, 2020) and second wave (December 16, 2020, through March 02, 2021) of the disease in SC. We measure the spatial (covariate-adjusted) INLA model at each time point using DIC (Deviance Information Criterion), Watanabe-Akaike Information Criterion (WAIC), and Log Marginal Likelihood. The minimum and maximum values of these diagnostics across time points are reported in the supporting information (**Table S1)**.

**Fig 1** displays the estimates of effective reproductive number for Charleston, Greenville, Horry, and Richland counties during the second COVID-19 wave in SC. In general, the proposed two-step spatial INLA estimates, 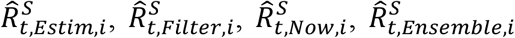, demonstrated reduced peaks compared to the initial estimates, 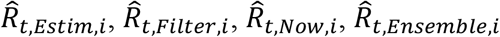.

**Fig. 1.**
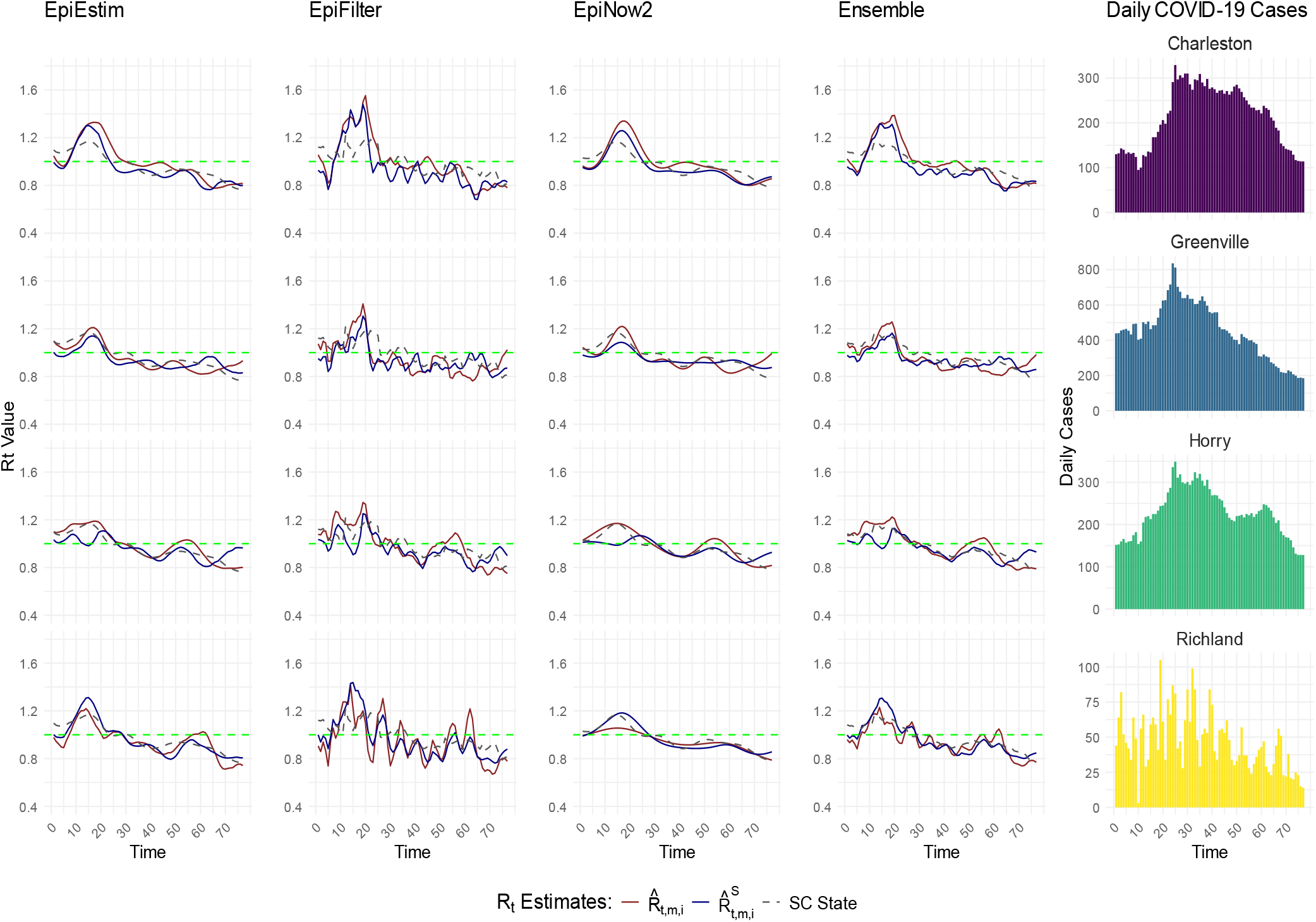
Comparison of two-step spatial (covariate-adjusted) INLA smoothing with initial *R*_*t*_ estimates at the county level in Wave 2. 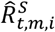 (blue line) represents the two-step spatial (covariate-adjusted) INLA smoothed estimates, 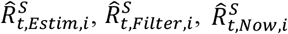, and 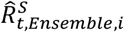, while 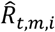 (red line) represents the initial estimates, 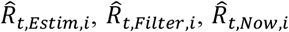, and 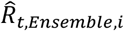, for select Charleston, Greenville, Horry, and Richland counties during COVID 19 Wave 2 (between December 16, 2020 – March 02, 2021) in SC. The state level initial estimate of *R*_*t*_ is presented with dashed gray line (SC State). The plots in the rightmost panel present the average daily cases for the respective counties over the same period.

**S1-S3 Figs** in the supplementary materials (Supporting Information 1) provide a detailed comparison of the effective reproductive number estimates, 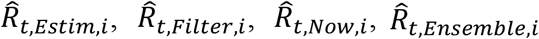 and 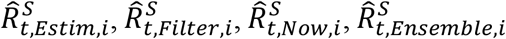, across various counties and ZIP codes. Results are presented for Charleston, Greenville, Horry, and Richland counties and ZIP codes 29605, 29642, 29680, and 29681, as these results serve as benchmarks when evaluating the prediction accuracy. The bar graphs displaying daily COVID-19 cases reveal spikes and declines that correspond to the fluctuations in the *R*_*t*_ estimates. For instance, peaks in case counts align with the increases in both sets of estimates, 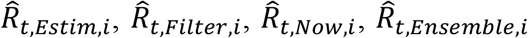 and 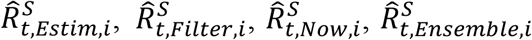. The figures show that when daily cases increase over a period, corresponding *R*_*t*_ estimates exceed 1.0, indicating infection growth. For example, in Fig.1, daily cases for Charleston, Greenville, and Horry counties increase between time points 10 and 25, with *R*_*t*_ estimates staying above 1.0. After time points 25, as cases decline almost everywhere, *R*_*t*_ estimates fall to 1.0 or lower, particularly with our two-step spatial INLA method (blue lines).

### Prediction of *R*_*t*_ in Regions with Entirely Missing Data

In this section, we evaluated the accuracy of our two-step spatial INLA model in estimating 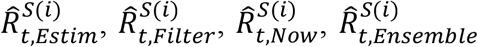 for regions with entirely missing data. We utilized daily COVID-19 case data at both county and ZIP code levels for the first wave (June 16, 2020, through August 31, 2020) and second wave (December 16, 2020, through March 02, 2021) of the pandemic in SC. **Table 1** presents the prediction accuracy achieved through the two-step spatial INLA model through comparison with the one-step estimates 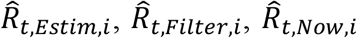, and 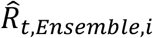. That is, we used the two-step spatial INLA model based on a subset of geographic regions (training set) to predict estimates of *R*_*t*_ for the remaining geographic region (i.e., validation set). Since there is no gold standard for comparison, we compared the predicted effective reproductive number, 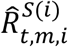, to the initial estimate 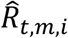.

**Table 1.**
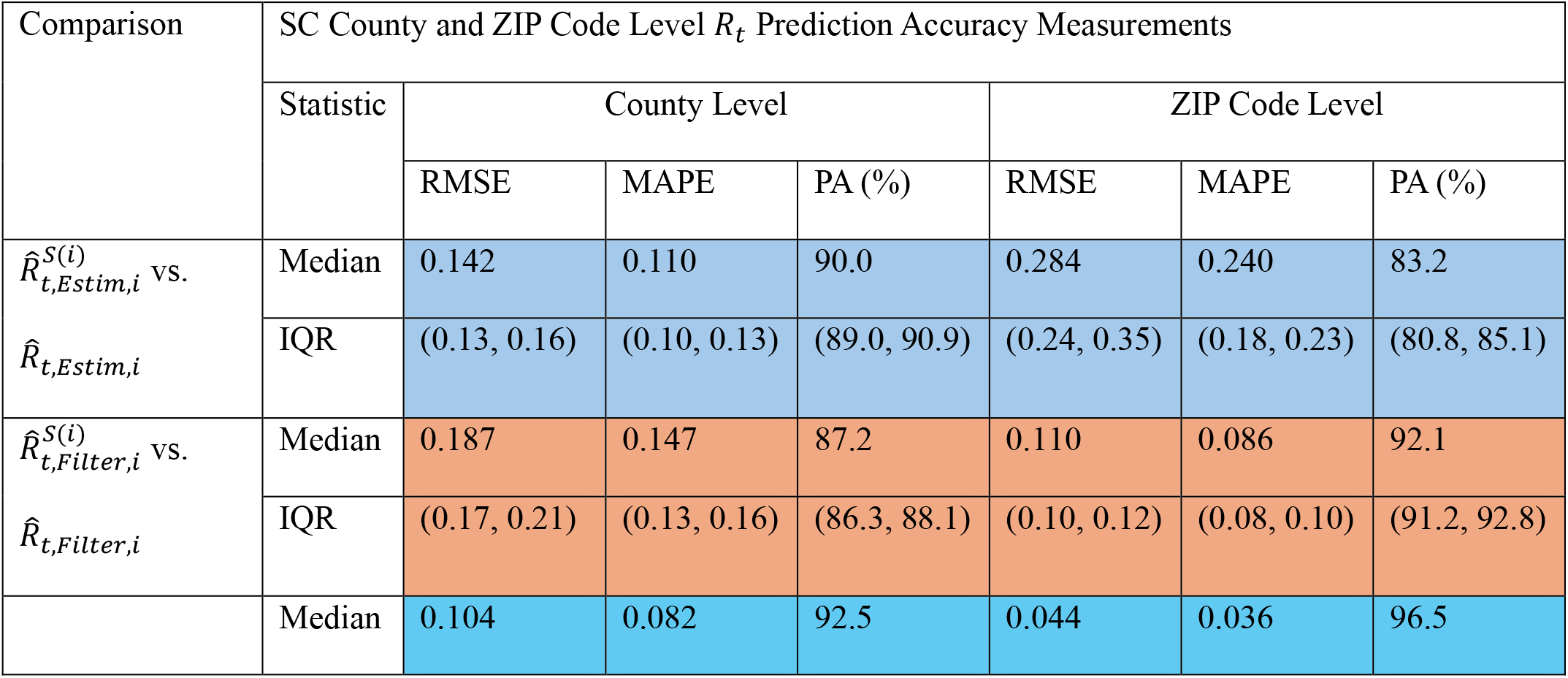
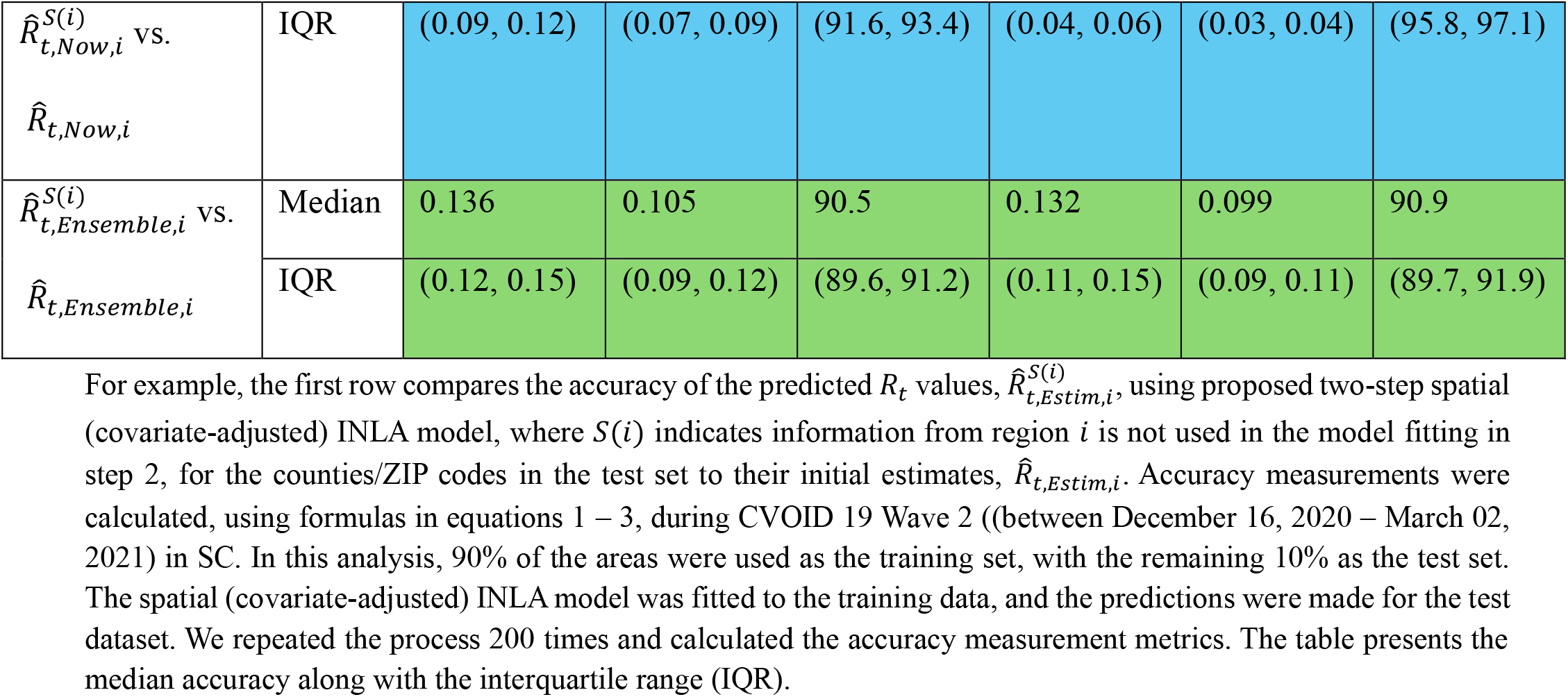
Comparison of two-step spatial (covariate-adjusted) INLA prediction, 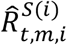, with initial estimates, 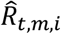, in Wave 2.

| Comparison | SC County and ZIP Code Level $R_t$ Prediction Accuracy Measurements | | | | | | |
| --- | --- | --- | --- | --- | --- | --- | --- |
|  | Statistic | County Level |  |  | ZIP Code Level |  |  |
|  |  | RMSE | MAPE | PA (%) | RMSE | MAPE | PA (%) |
| $\hat{R}_{t,Estim,i}^{S(i)}$ vs.<br>$\hat{R}_{t,Estim,i}$ | Median | 0.142 | 0.110 | 90.0 | 0.284 | 0.240 | 83.2 |
|  | IQR | (0.13, 0.16) | (0.10, 0.13) | (89.0, 90.9) | (0.24, 0.35) | (0.18, 0.23) | (80.8, 85.1) |
| $\hat{R}_{t,Filter,i}^{S(i)}$ vs.<br>$\hat{R}_{t,Filter,i}$ | Median | 0.187 | 0.147 | 87.2 | 0.110 | 0.086 | 92.1 |
|  | IQR | (0.17, 0.21) | (0.13, 0.16) | (86.3, 88.1) | (0.10, 0.12) | (0.08, 0.10) | (91.2, 92.8) |
|  | Median | 0.104 | 0.082 | 92.5 | 0.044 | 0.036 | 96.5 |
| $\hat{R}_{t,Now,i}^{S(i)}$ vs.<br>$\hat{R}_{t,Now,i}$ | IQR | (0.09, 0.12) | (0.07, 0.09) | (91.6, 93.4) | (0.04, 0.06) | (0.03, 0.04) | (95.8, 97.1) |
| $\hat{R}_{t,Ensemble,i}^{S(i)}$ vs.<br>$\hat{R}_{t,Ensemble,i}$ | Median | 0.136 | 0.105 | 90.5 | 0.132 | 0.099 | 90.9 |
|  | IQR | (0.12, 0.15) | (0.09, 0.12) | (89.6, 91.2) | (0.11, 0.15) | (0.09, 0.11) | (89.7, 91.9) |

For example, the first row compares the accuracy of the predicted $R_t$ values, $\hat{R}_{t,Estim,i}^{S(i)}$ , using proposed two-step spatial (covariate-adjusted) INLA model, where $S(i)$ indicates information from region $i$ is not used in the model fitting in step 2, for the counties/ZIP codes in the test set to their initial estimates, $\hat{R}_{t,Estim,i}$ . Accuracy measurements were calculated, using formulas in equations 1 – 3, during CVOID 19 Wave 2 ((between December 16, 2020 – March 02, 2021) in SC. In this analysis, 90% of the areas were used as the training set, with the remaining 10% as the test set. The spatial (covariate-adjusted) INLA model was fitted to the training data, and the predictions were made for the test dataset. We repeated the process 200 times and calculated the accuracy measurement metrics. The table presents the median accuracy along with the interquartile range (IQR).

At both county and ZIP code levels during the second wave of COVID-19, predicted *R*_*t*_ values, 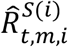, based on the two-step spatial INLA estimators achieved high level of accuracy (**Table 1**).

For counties, the median percentage agreement was 90.0% (IQR: 89.0–90.9 %) for 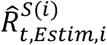, 87.2% (IQR: 86.3–88.1 %) for 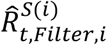, 92.5% (IQR: 91.6–93.4 %) for 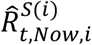, and 90.5% (IQR: 89.6–91.2 %) for 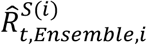. The prediction, 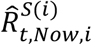, achieved the highest accuracy, with a median PA of 92.5% at the county level. Similarly, 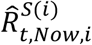 exhibited the highest median PA of 96.5% for SC ZIP codes. In the first wave, the accuracy was also high, particularly for 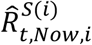, which obtained a median PA of 90.9% at the county level and 95.2% at the ZIP code level (**see Table S2**). However, 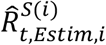 showed the lowest accuracy, with a median PA of 81.9% at the ZIP code level, and a median PA of 85.7% at the county level. Overall, the two-step spatial (covariate-adjusted) INLA procedure showed high performance accuracy in computing 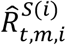 for areas with missing data, as further evaluated through root mean squared error (RMSE) and mean absolute percentage error (MAPE).

In the second validation approach, we selected Charleston, Greenville, Horry, and Richland counties (county level analysis), and ZIP codes 29605, 29642, 29680, and 29681 (zip code level analysis) as the test set. **Fig 2** provides a detailed comparison of the predictions 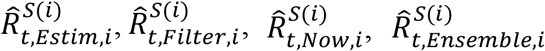 to initial estimates 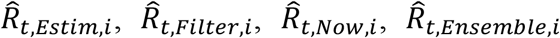 across counties–Charleston, Greenville, Horry, and Richland and ZIP codes– 29605, 29642, 29680, and 29681 (also see **S4-S8 Figs**). The results comparing prediction accuracy for this validation setting are presented in **Table S3** for the first wave and **Table S4** for the second wave. During the second wave, 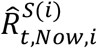 demonstrated the highest accuracy, with a percentage agreement (PA) range of 91.1% - 94.0% across the four counties and 96.1% – 98.7 % across the four ZIP codes. 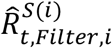 achieved the lowest PA range for counties (86.7% – 90.8%) and 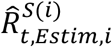 achieved the lowest PA range for ZIP codes (86.9%–91.5 %). Similar results were obtained during the first wave.

**Fig. 2.**
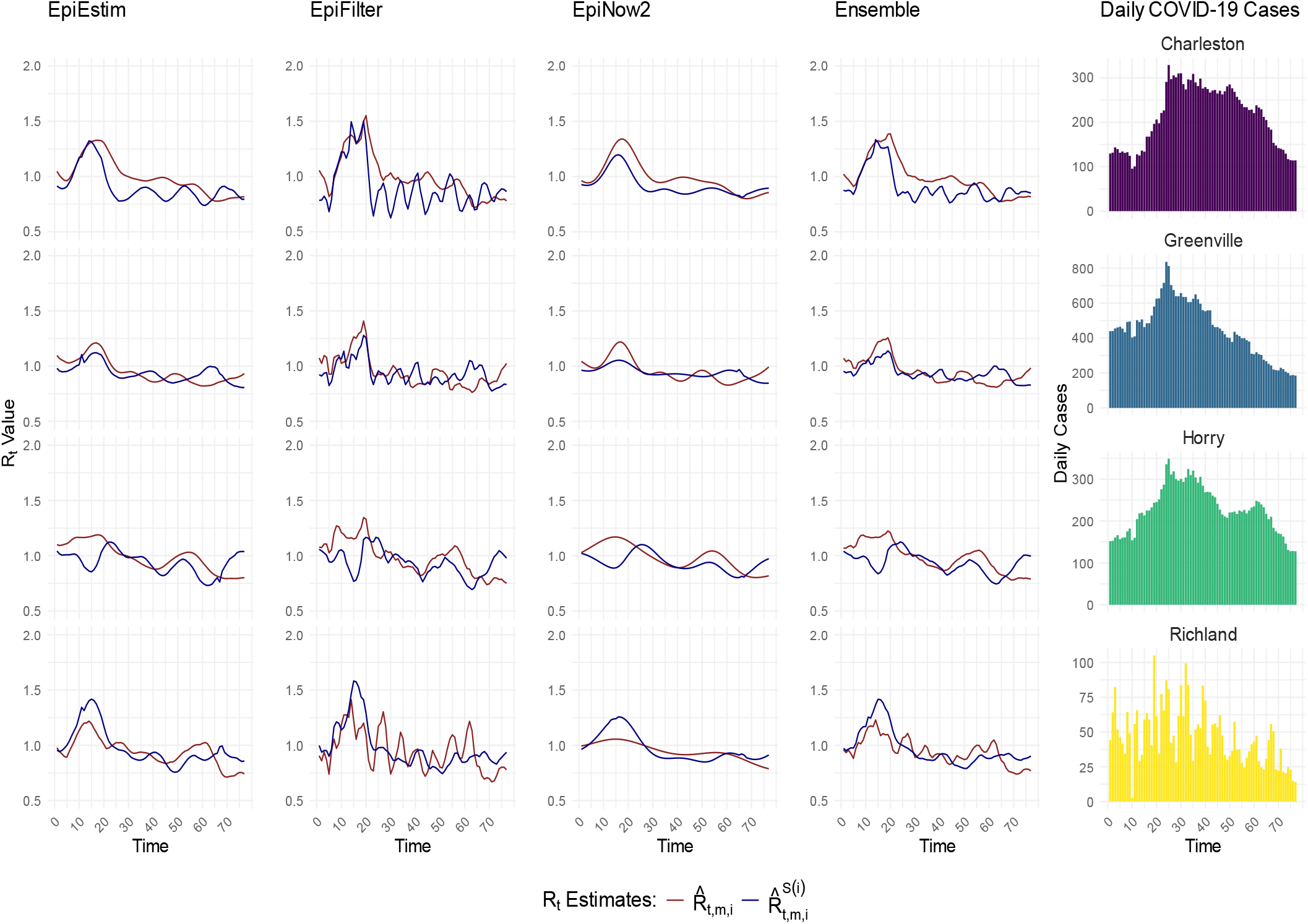
Comparison of two-step spatial (covariate-adjusted) INLA prediction with the initial *R*_*t*_ estimates in Wave 2. 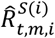 (blue line) represents the two-step spatial (covariate-adjusted) INLA prediction of effective reproductive number, 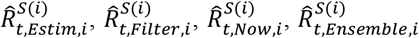, while 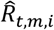 (red line) represents the initial estimates, 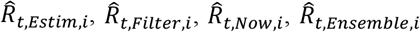, for select Charleston, Greenville, Horry, and Richland counties during COVID 19 Wave 2 (between December 16, 2020 – March 02, 2021) in SC. Here, *S*(*i*) *in* 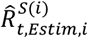 indicates that geographic region *i* was not included in the INLA model fitting. For example, if 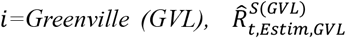 means Greenville County was not used in the spatial (covariate-adjusted) INLA model fitting in step 2. The plots in the rightmost panel present the average daily cases for the respective counties over the same period.

### Simulation Results

We evaluated the performance of our two-step spatial (covariate-adjusted) INLA smoothing and prediction framework for estimating *R*_*t*_ at the county level. The analysis was conducted using simulated COVID-19 case counts data for counties in SC and focused on assessing the accuracy of initial estimates, improvements achieved through spatial INLA smoothing, and predictive ability of the framework for counties with completely missing data.

Figure **S9** presents a comparison between the initial *R*_*t*_ estimates obtained using EpiEstim, EpiFilter, and the ensemble approach, the smoothed *R*_*t*_ estimates generated through the two-step spatial (covariate-adjusted) INLA smoothing, and the true generated *R*_*t*_ values used in the simulation. The results demonstrate that the initial estimates provide a reasonable estimation of *R*_*t*_ across counties. However, after applying spatial INLA smoothing, the estimates align more closely with the true generated *R*_*t*_ values. The percentage agreement (PA) values in Table **S6** also indicate that the smoothed *R*_*t*_ estimates align well with the true values. In this scenario, all available data were used, meaning that the initial estimates were available for all counties before applying the spatial INLA model.

Figure **S10** evaluates the predictive capability of our approach for estimating *R*_*t*_ in counties with completely missing data. To simulate this scenario, we excluded each county one by one from the model fitting process. For example, Charleston County was left out to replicate a situation where no data was available for that county. The model then predicted *R*_*t*_ for Charleston by incorporating spatial information from neighboring counties along with sociodemographic covariate data specific to the county. This process was repeated for Greenville, Horry, and Richland counties to evaluate the model’s robustness. The results show that the *R*_*t*_ values predicted using our two-step spatial (covariate-adjusted) INLA model closely align with the true generated *R*_*t*_ values. The percentage agreement values in Table **S6** further validate the robustness of the proposed framework, showing that it maintains high prediction accuracy even for the counties with missing data.

For the initial estimates, the median PA was 92.55% (IQR: 92.63%-93.74%) for 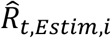, 91.52% (IQR: 91.12-91.83%) for 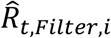, and 92.09% (IQR: 91.72-92.37%) for 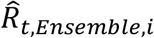. Our framework showed higher performance, the PA was 93.10% (IQR: 92.43-93.75%) for 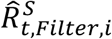 and 93.10% (IQR: 92.47-93.91%) for 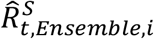. For region with entirely missing data that makes impossible to estimate *R*_*t*_ with the existing methods, the PA was 90.76% (IQR: 89.67-91.38%) for 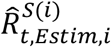, 91.30% (IQR: 90.79-91.57%) for 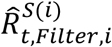. These findings highlight the effectiveness of our two-step spatial (covariate-adjusted) INLA framework in estimating *R*_*t*_ at the local level with coarse or missing data.

## Discussion

Estimation of time-varying effective reproductive number *R*_*t*_ is necessary for understanding the transmission dynamics of infectious disease and implementing the necessary public health interventions. We developed a two-step procedure for estimating the effective reproductive number for infectious diseases in granular geographic regions. Our approach incorporates existing *R*_*t*_ estimation procedures (EpiEstim, EpiFilter, and EpiNow2), along with an ensemble-based estimate using data from geographic regions with sufficient data (step 1), into a spatial modeling framework to predict *R*_*t*_ in regions with sparse or missing data (step 2). The choice of *R*_*t*_ estimation methods is not exhaustive. Our flexible framework allows us to implement any existing estimation procedure for *R*_*t*_ in regions with coarse or entirely missing data.

Our proposed methodology improves the estimation of the effective reproductive number *R*_*t*_ in small areas, especially in areas where the disease data are coarse or missing. By integrating the Bayesian spatial INLA model, this method efficiently utilizes the available data and borrows information from neighboring areas and similar regions through sociodemographic covariates, enhancing the robustness of *R*_*t*_ estimates. Decisions regarding the implementation, removal, or adjustment of infection control measures are guided by a combination of epidemiological, social, and economic factors [36,37]. Effective reproductive number estimates assist in this process by providing critical information about the potential future course of an outbreak. Through small area estimation of the effective reproductive number, we can better identify and prioritize high risk areas, which allows for optimal resource allocation [2]. For example, this current NIH and CDC-funded study is intended to better identify and prioritize medically underserved communities at high-risk of infectious disease outbreaks for priority delivery of mobile health clinics for infectious disease screening, treatment, and vaccination.

Our study has several limitations. One primary limitation is that our approach focuses on the retroactive estimation of *R*_*t*_. This is particularly useful for understanding past outbreaks and for forecasting outbreaks when the effective reproductive number in each region is similar between outbreaks (e.g., first two waves of Covid-19 in SC; **Table S5**). While this can be a reasonable assumption for endemic viruses, this is far from guaranteed for emerging viruses where disease transmission dynamics and/or human behavior changes can substantially change with the introduction of a new variant. Therefore, a natural extension is to extend our approach for forecasting future values of *R*_*t*_, particularly in granular geographic regions with sparse data.

We evaluated model performance using three commonly used techniques (EpiEstim, EpiFilter, EpiNow2) along with the ensemble-based estimation. Similar to most *R*_*t*_ estimation methods, EpiEstim and EpiFilter rely on the reported incidence data [6,17]. Inaccuracies or delays in reporting can lead to biased estimates. EpiNow2 also makes certain assumptions about the underlying transmission dynamics, such as the generation time distribution and reporting delays. These assumptions can introduce uncertainty into the estimates, particularly during the periods of low incidence [18]. Additionally, our study did not include the other *R*_*t*_ estimation procedures, such as Epidemia [38], EpiInvert [39], EpiRegress [40], estimateR [41], GrowthPredict[42], and Extended Kalman Filter [43].

Due to the limitations of using existing *R*_*t*_ estimators as a benchmark to evaluate the performance of our proposed estimator, there is no gold standard for comparison. To address this, we performed a simulation study to compare the *R*_*t*_ estimates with ground truth *R*_*t*_ values. While the simulation study demonstrated our proposed estimator was accurate and closely resembled ground truth estimates, it was conducted in a single simulation setting, with fixed parameter ranges and temporal dynamics, which may not fully capture the diversity of real-world scenarios.

Future research may extend our framework by incorporating the forecasting of *R*_*t*_ values and subsequent case projections, providing critical insights into future disease dynamics. This would help policymakers with decision-making, support outbreak detection, and strengthen preparedness for future outbreaks. In our current approach, we utilized three existing *R*_*t*_estimation methods and combined them using an ensemble approach with equal weights. Future effort also could explore incorporating additional estimation methods and developing an ensemble estimation technique with optimal weights to improve the accuracy and robustness across various scenarios. Furthermore, simulations under different settings could be conducted to capture a wider range of epidemic scenarios.

## Conclusion

This study presented a two-step spatial INLA approach to estimate the effective reproductive number in small areas with coarse or missing data. Our approach incorporated existing estimators into a Bayesian spatial model that adjusted for spatial variation by borrowing information from neighboring regions and incorporating sociodemographic covariates. This approach provided predictions of the effective reproductive number for regions with missing data. Importantly, our proposed framework is flexible, allowing for implementation of any existing estimation procedure for *R*_*t*_ in geographically granular regions. While this study focused on Covid-19, our methodology is translatable for estimation of the effective reproductive number for other respiratory infectious diseases.

## Data Availability

All data produced in the present study are available upon reasonable request to the authors.

## Supporting Information 1

**S1 Table**. INLA model diagnostics (DIC, WAIC, and Log Marginal Likelihood) for Wave 1 and Wave 2.

**S1 Fig**. Comparison of two-step spatial (covariate-adjusted) INLA smoothing with initial *R*_*t*_ estimates at the county level in Wave 1.

**S2 Fig**. Comparison of two-step spatial (covariate-adjusted) INLA smoothing with initial *R*_*t*_ estimates at the ZIP code level in Wave 1.

**S3 Fig**. Comparison of two-step spatial (covariate-adjusted) INLA smoothing with initial *R*_*t*_ estimates at the ZIP code level in Wave 2.

**S2 Table**. Comparison of two-step spatial (covariate-adjusted) INLA prediction, 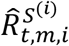, with initial estimates, 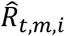, in Wave 1.

**S3 Table**. Comparison (leave-one-out validation) of two-step spatial (covariate-adjusted) INLA prediction, 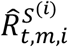, with initial estimates, 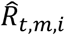, in Wave 1.

**S4 Table**. Comparison (leave-one-out validation) of two-step spatial (covariate-adjusted) INLA prediction, 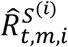, with initial estimates, 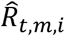, in Wave 2.

**S4 Fig**. Comparison of two-step spatial (covariate-adjusted) INLA prediction with the initial *R*_*t*_ estimates at the county level in Wave 1.

**S5 Fig**. Comparison of two-step spatial (covariate-adjusted) INLA prediction with the initial *R*_*t*_ estimates at the ZIP code level in Wave 1.

**S6 Fig**. Comparison of two-step spatial (covariate-adjusted) INLA prediction with the initial *R*_*t*_ estimates at the ZIP code level in Wave 2.

**S7 Fig**. Comparison of *R*_*t*_ estimates of the county Greenville and its neighboring counties in Wave 2.

**S8 Fig**. Comparison of *R*_*t*_ estimates of the county Greenville and its neighboring counties in Wave 2.

**S5 Table**. Comparison of initial estimates and two-step INLA spatial (covariate-adjusted) estimates between Wave 1 and Wave 2.

**S9 Fig**. Comparison of two-step spatial (covariate-adjusted) INLA smoothing and initial *R*_*t*_ estimates with the generated true *R*_*t*_.

**S10 Fig**. Comparison of two-step spatial (covariate-adjusted) INLA prediction with the generated true *R*_*t*_.

**S6 Table**. Comparison of initial estimates, two-step spatial (covariate-adjusted) INLA smoothed estimates, and predicted *R*_*t*_ with the true generated *R*_*t*_.

## Abbreviations

INLA: Integrated Nested Laplace Approximation
SC: South Carolina
PA: Percentage Agreement
IQR: Interquartile Range
NIH: National Institutes of Health
CDC: Center for Disease Control and Prevention
SVI: Social Vulnerability Index
NYT: New York Times
SD: Standard Deviation
CI: Confidence Interval
RMSE: Root Mean Squared Error
MAPE: Mean Absolute Percentage Error
GVL: Greenville
DIC: Deviance Information Criterion
WAIC: Watanabe-Akaike Information Criterion

## Acknowledgements

Thanks to Tanvir Ahammed and Dr. Jiande Wu for their assistance with processing COVID-19 case data at the ZIP code and county levels in SC.

## Author Contributions

**Conceptualization:** MSH, RG, NKM, VDG, CM, LR

**Formal Analysis:** MSH

**Investigation:** MSH, LR

**Methodology:** MSH, RV, NKM, VDG, CM, LR

**Project administration:** LR

**Software:** MSH

**Supervision:** LR, CM

**Validation:** MSH

**Visualization:** MSH

**Writing-Original Draft Preparation:** MSH, LR

**Writing-Review & Editing:** MSH, RG, NKM, VDG, MMC, CM, LR

## Data Availability

All code and public data used for analyses and figure generations are available at https://github.com/mdsakhh/Small-Area-Estimation-of-Effective-Reproductive-Number and from the corresponding author upon reasonable request.

## Declarations

### Ethics approval and consent to participate

Ethical review for this study was obtained by the Institutional Review Board of Clemson University (#2020-0150). No consent was needed for this study; retrospective data is based on medical claims and electronic health records and were de-identified to study investigators.

### Clinical Trial Number

Not applicable.

### Consent for publication

Not applicable.

