## Supplementary Information 1 for "A Flexible Framework for Local-Level Estimation of the Effective Reproductive Number in Geographic Regions with Sparse Data": supporting_information.pdf

### Supporting Information 1

#### Supplementary Figures and Tables

**S1 Table.** INLA model diagnostics (DIC, WAIC, and Log Marginal Likelihood) for Wave 1 and Wave 2.

| Method | Data | County | | | ZIP Code | | |
|---|---|---|---|---|---|---|---|
| | | DIC (Min, Max) | WAIC (Min, Max) | Log Marginal Likelihood (Min, Max) | DIC (Min, Max) | WAIC (Min, Max) | Log Marginal Likelihood (Min, Max) |
| EpiEstim | Wave 1 | (-249.0, 89.4) | (-258.0, 98.3) | (-102.0, -40.9) | (-162.0, 43.0) | (-168.0, 44.2) | (-62.0, -31.6) |
| | Wave 2 | (-257.0, -12.3) | (-267.0, -7.53) | (-61.2, -25.7) | (-36.2, 75.9) | (-34.0, 80.8) | (-88.5, -44.7) |
| EpiFilter | Wave 1 | (-248.0, 26.0) | (-257.0, 26.4) | (-75.8, -39.5) | (-165.0, -16.3) | (-171.0, -16.7) | (-40.8, -15.7) |
| | Wave 2 | (-241.0, 26.0) | (-248.0, 29.8) | (-76.0, -37.6) | (-130.0, -28.3) | (-128.0, -28.2) | (-47.8, -8.41) |
| EpiNow2 | Wave 1 | (-251.0, -10.9) | (-261.0, -9.45) | (-61.6, -26.3) | (-166.0, -49.3) | (-167.0, -49.5) | (-29.2, -8.72) |
| | Wave 2 | (-241.0, -37.0) | (-248.0, -33.4) | (-51.7, -15.6) | (-214.0, -116.0) | (-211.0, -113.0) | (-13.9, 22.1) |
| Ensemble | Wave 1 | (-250.0, 32.1) | (-260.0, 37.0) | (-78.3, -36.5) | (-158.0, -5.45) | (-163.0, -4.11) | (-44.8, -20.3) |
| | Wave 2 | (-252.0, -9.30) | (-262.0, -4.56) | (-62.3, -27.0) | (-97.0, -3.65) | (-95.8, 0.014) | (-57.4, -21.0) |

This table presents the Deviance Information Criterion (DIC), Watanabe-Akaike Information Criterion (WAIC), and Log Marginal Likelihood as model diagnostics for Bayesian spatial (covariate-adjusted) INLA model models. These diagnostics are calculated for two distinct COVID-19 waves: Wave 1 (June 16, 2020 – August 31, 2020) and Wave 2 (December 16, 2020 – March 2, 2021), at both the county and ZIP code levels. The models were fitted separately at each time point to perform spatial covariate-adjusted smoothing of $R_t$ estimates. The minimum and maximum DIC, WAIC, and Log Marginal Likelihood values at county and ZIP code levels confirm the models' convergence across time points.

#### Figures S1-S3:

**Comparison of the effective reproductive number estimates $\hat{R}^S_{t,Estim,i}$, $\hat{R}^S_{t,Filter,i}$, $\hat{R}^S_{t,Now,i}$, $\hat{R}^S_{t,Ensemble,i}$ with $\hat{R}_{t,Estim,i}$, $\hat{R}_{t,Filter,i}$, $\hat{R}_{t,Now,i}$, $\hat{R}_{t,Ensemble,i}$ at the county and ZIP code levels in SC.**

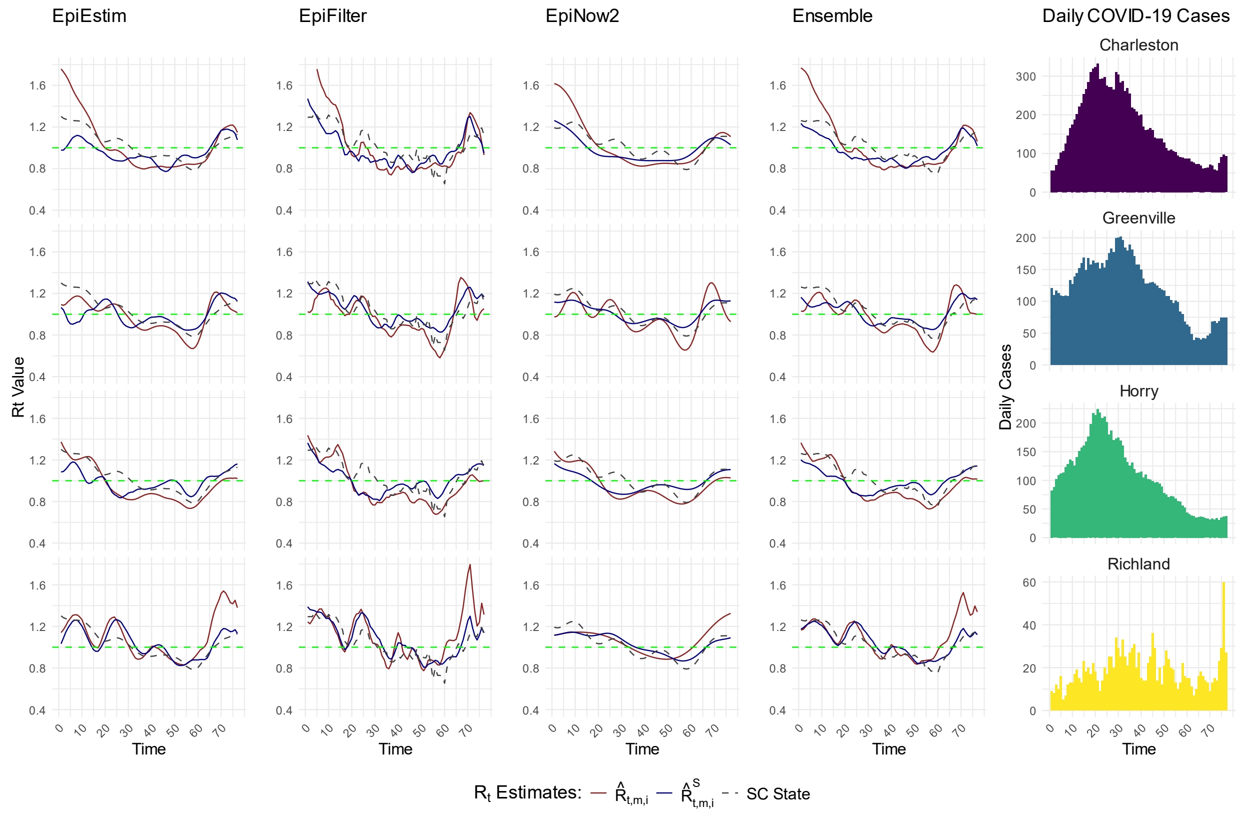


**S1 Fig.** Comparison of two-step spatial (covariate-adjusted) INLA smoothing with initial $R_t$ estimates at the county level in Wave 1.

$\hat{R}^S_{t,m,i}$ (blue line) represents the two-step spatial (covariate-adjusted) INLA smoothed estimates, $\hat{R}^S_{t,Estim,i}$, $\hat{R}^S_{t,Filter,i}$, $\hat{R}^S_{t,Now,i}$, and $\hat{R}^S_{t,Ensemble,i}$, while $\hat{R}_{t,m,i}$ (red line) represents the initial estimates, $\hat{R}_{t,Estim,i}$, $\hat{R}_{t,Filter,i}$, $\hat{R}_{t,Now,i}$, and $\hat{R}_{t,Ensemble,i}$, for select Charleston, Greenville, Horry, and Richland counties during COVID 19 Wave 1 (between June 16, 2020 – August 31, 2020) in SC. The state level initial estimate of $R_t$ is presented with dashed gray line (SC State). The plots in the rightmost panel present the average daily cases for the respective counties over the same period.

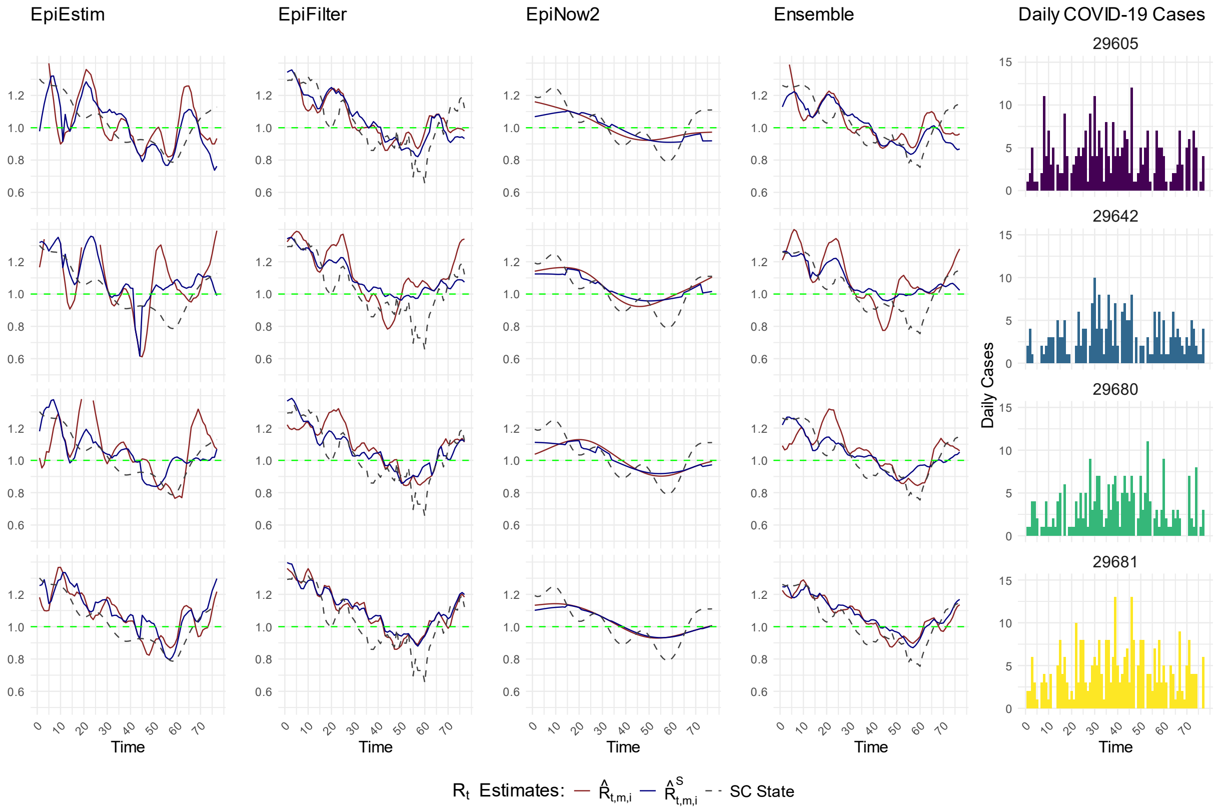


**S2 Fig.** Comparison of two-step spatial (covariate-adjusted) INLA smoothing with initial $R_t$ estimates at the ZIP code level in Wave 1.

$\hat{R}^{S}_{t,m,i}$ (blue line) represents the two-step spatial (covariate-adjusted) INLA smoothed estimates, $\hat{R}^{S}_{t,Estim,i}$, $\hat{R}^{S}_{t,Filter,i}$, $\hat{R}^{S}_{t,Now,i}$, and $\hat{R}^{S}_{t,Ensemble,i}$, while $\hat{R}_{t,m,i}$ (red line) represents the initial estimates, $\hat{R}_{t,Estim,i}$, $\hat{R}_{t,Filter,i}$, $\hat{R}_{t,Now,i}$, and $\hat{R}_{t,Ensemble,i}$, for select- 29605, 29642, 29680, and 29681, ZIP codes during COVID 19 Wave 1 (between June 16, 2020 – August 31, 2020) in SC. The state level initial estimate of $R_t$ is presented with dashed gray line (SC State). The plots in the right most panel present the average daily cases for the respective ZIP codes over the same period.

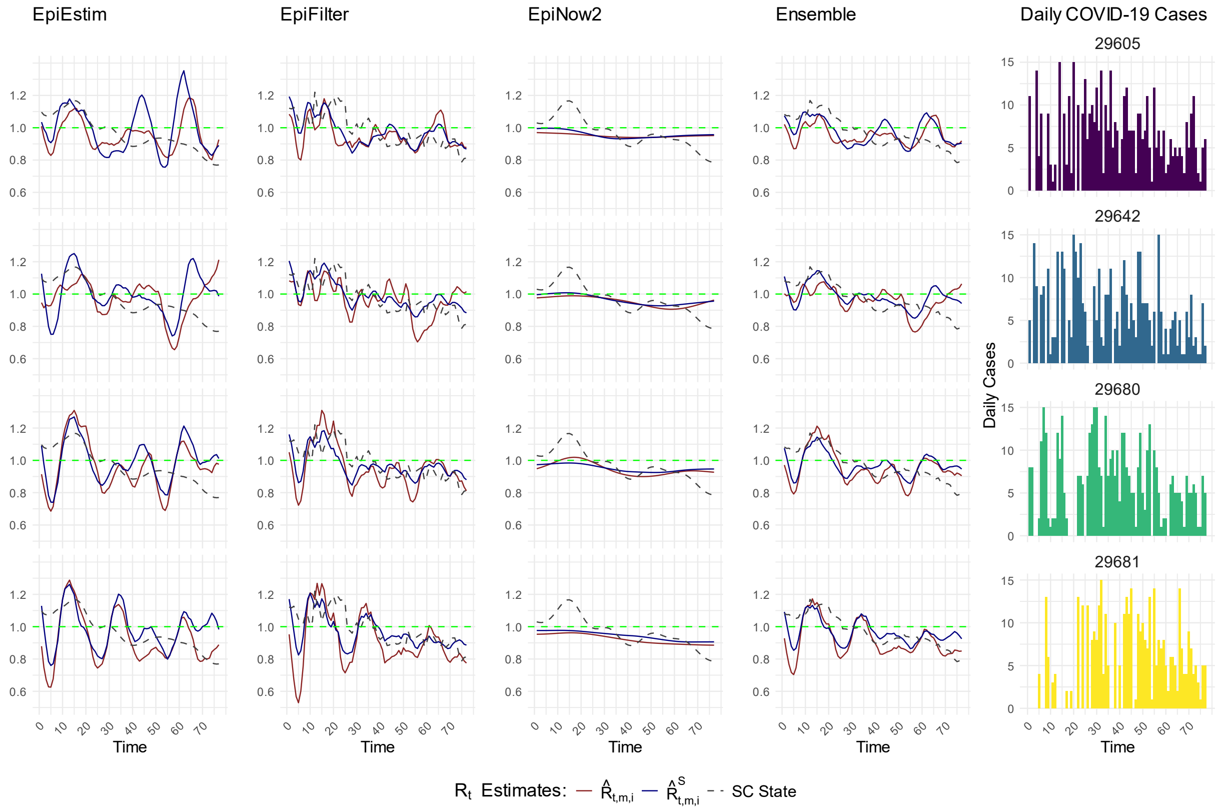


**S3 Fig.** Comparison of two-step spatial (covariate-adjusted) INLA smoothing with initial $R_t$ estimates at the ZIP code level in Wave 2.

$\hat{R}^S_{t,m,i}$ (blue line) represents the two-step spatial (covariate-adjusted) INLA smoothed estimates, $\hat{R}^S_{t,Estim,i}$, $\hat{R}^S_{t,Filter,i}$, $\hat{R}^S_{t,Now,i}$, and $\hat{R}^S_{t,Ensemble,i}$, while $\hat{R}_{t,m,i}$ (red line) represents the initial estimates, $\hat{R}_{t,Estim,i}$, $\hat{R}_{t,Filter,i}$, and $\hat{R}_{t,Now,i}$, for select- 29605, 29642, 29680, and 29681 ZIP codes during COVID 19 Wave 2 (between December 16, 2020 – March 02, 2021) in SC. The state level initial estimate of $R_t$ is presented with dashed gray line (SC State). The plots in the right most panel present the average daily cases for the respective ZIP codes over the same period.

**Tables S2, S3, S4 and Figures S4, S5, S6:**

**Comparison of two-step spatial (covariate-adjusted) INLA prediction $\hat{R}^{S(i)}_{t,m,i}$ with initial estimates $\hat{R}_{t,m,i}$ at the county and ZIP code levels in SC.**

**S2 Table.** Comparison of two-step spatial (covariate-adjusted) INLA prediction, $\hat{R}^{S(i)}_{t,m,i}$, with initial estimates, $\hat{R}_{t,m,i}$, in Wave 1.

| Comparison | SC County and ZIP Code Level $R_t$ Prediction Accuracy Measurements | | | | | | |
|---|---|---|---|---|---|---|---|
| | Statistic | County Level | | | ZIP Code Level | | |
| | | RMSE | MAPE | PA (%) | RMSE | MAPE | PA (%) |
| $\hat{R}^{S(i)}_{t,Estim,i}$ vs. $\hat{R}_{t,Estim,i}$ | Median | 0.265 | 0.167 | 85.7 | 0.337 | 0.218 | 81.9 |
| | IQR | (0.23, 0.35) | (0.15, 0.19) | (84.4, 87.1) | (0.30, 0.41) | (0.20, 0.25) | (79.6, 83.5) |
| $\hat{R}^{S(i)}_{t,Filter,i}$ vs. $\hat{R}_{t,Filter,i}$ | Median | 0.206 | 0.145 | 87.4 | 0.134 | 0.096 | 91.3 |
| | IQR | (0.18, 0.23) | (0.13, 0.16) | (86.2, 88.0) | (0.12, 0.16) | (0.08, 0.11) | (89.7, 92.2) |
| $\hat{R}^{S(i)}_{t,Now,i}$ vs. $\hat{R}_{t,Now,i}$ | Median | 0.137 | 0.099 | 90.9 | 0.067 | 0.050 | 95.2 |
| | IQR | (0.11, 0.17) | (0.09, 0.11) | (89.9, 92.0) | (0.06, 0.08) | (0.04, 0.06) | (94.4, 95.7) |
| $\hat{R}^{S(i)}_{t,Ensemble,i}$ vs. $\hat{R}_{t,Ensemble,i}$ | Median | 0.192 | 0.131 | 88.4 | 0.161 | 0.110 | 90.0 |
| | IQR | (0.16, 0.23) | (0.12, 0.15) | (87.4, 89.4) | (0.14, 0.19) | (0.10, 0.13) | (88.5, 90.9) |

For example, the first row compares the accuracy of the predicted $R_t$ values, $\hat{R}^{S(i)}_{t,Estim,i}$, using proposed two-step spatial (covariate-adjusted) INLA model, where $S(i)$ indicates information from region $i$ is not used in the model fitting in step 2, of the counties/ZIP codes from the test set to their initial estimates, $\hat{R}_{t,Estim,i}$. Accuracy measurements were calculated, using formulas in equations 1 – 3, during CVOID 19 Wave 1 (between June 16, 2020 – August 31, 2020) in SC. In this analysis, 90% of the areas were used as the training set, with the remaining 10% as the test set. The spatial (covariate-adjusted) INLA model was fitted to the training data, and the predictions were made for the test data set. We repeated the process 200 times and calculated the accuracy measurement metrics. The table presents the median accuracy along with the interquartile range (IQR).

**S3 Table.** Comparison (leave-one-out validation) of two-step spatial (covariate-adjusted) INLA prediction, $\hat{R}_{t,m,i}^{S(i)}$, with initial estimates, $\hat{R}_{t,m,i}$, in Wave 1.

| Comparison | SC County and ZIP Code Level $R_t$ Prediction Accuracy Measurements | | | | | | |
|---|---|---|---|---|---|---|---|
| | Statistic | County Level | | | ZIP Code Level | | |
| | | RMSE | MAPE | PA (%) | RMSE | MAPE | PA (%) |
| $\hat{R}_{t,Estim,i}^{S(i)}$ vs. $\hat{R}_{t,Estim,i}$ | Median | 0.203 | 0.148 | 86.6 | 0.223 | 0.157 | 86.1 |
| | Range | (0.14, 0.39) | (0.12, 0.20) | (80.7, 88.9) | (0.21, 0.23) | (0.14, 0.19) | (83.4, 87.0) |
| $\hat{R}_{t,Filter,i}^{S(i)}$ vs. $\hat{R}_{t,Filter,i}$ | Median | 0.206 | 0.162 | 86.3 | 0.105 | 0.079 | 92.4 |
| | Range | (0.15, 0.33) | (0.13, 0.19) | (82.0, 88.1) | (0.09, 0.12) | (0.06, 0.09) | (91.7, 94.1) |
| $\hat{R}_{t,Now,i}^{S(i)}$ vs. $\hat{R}_{t,Now,i}$ | Median | 0.138 | 0.125 | 89.0 | 0.043 | 0.033 | 96.7 |
| | Range | (0.13, 0.25) | (0.08, 0.14) | (86.0, 91.8) | (0.03, 0.06) | (0.02, 0.05) | (94.9, 98.3) |
| $\hat{R}_{t,Ensemble,i}^{S(i)}$ vs. $\hat{R}_{t,Ensemble,i}$ | Median | 0.176 | 0.140 | 88.0 | 0.118 | 0.080 | 92.3 |
| | Range | (0.12, 0.33) | (0.10, 0.18) | (83.0, 90.3) | (0.10, 0.12) | (0.07,0.10) | (90.7, 92.9) |

Comparison of two-step spatial (covariate-adjusted) INLA prediction, $\hat{R}_{t,m,i}^{S(i)}$, with initial estimates, $\hat{R}_{t,m,i}$, using the leave-one-out validation approach during Wave 1 (between June 16, 2020 – August 31, 2020). The prediction accuracy is presented for counties–Charleston, Greenville, Horry, and Richland and ZIP codes – 29605, 29642, 29680, and 29681. For example, the first row compares the accuracy of the predicted $R_t$ values, $\hat{R}_{t,Estim,i}^{S(i)}$, using proposed two-step spatial (covariate-adjusted) INLA model, where $S(i)$ indicates information from region $i$ is not used in the model fitting in step 2, to their initial estimates, $\hat{R}_{t,Estim,i}$, for these selected counties and ZIP codes.

**S4 Table.** Comparison (leave-one-out validation) of two-step spatial (covariate-adjusted) INLA prediction, $\hat{R}^{S(i)}_{t,m,i}$, with initial estimates, $\hat{R}_{t,m,i}$, in Wave 2.

| Comparison | SC County and ZIP Code Level $R_t$ Prediction Accuracy Measurements | | | | | | |
|---|---|---|---|---|---|---|---|
| | Statistic | County Level | | | ZIP Code Level | | |
| | | RMSE | MAPE | PA (%) | RMSE | MAPE | PA (%) |
| $\hat{R}^{S(i)}_{t,Estim,i}$ vs. | Median | 0.129 | 0.109 | 89.7 | 0.150 | 0.123 | 89.2 |
| $\hat{R}_{t,Estim,i}$ | Range | (0.08, 0.14) | (0.08, 0.11) | (89.2, 92.8) | (0.11, 0.19) | (0.09, 0.17) | (86.9, 91.5) |
| $\hat{R}^{S(i)}_{t,Filter,i}$ vs. | Median | 0.166 | 0.132 | 87.5 | 0.095 | 0.083 | 92.5 |
| $\hat{R}_{t,Filter,i}$ | Range | (0.11, 0.18) | (0.10, 0.15) | (86.7, 90.8) | (0.07, 0.19) | (0.06, 0.19) | (85.7, 94.7) |
| $\hat{R}^{S(i)}_{t,Now,i}$ vs. | Median | 0.093 | 0.074 | 92.8 | 0.023 | 0.020 | 98.0 |
| $\hat{R}_{t,Now,i}$ | Range | (0.08, 0.12) | (0.06, 0.09) | (91.1, 94.0) | (0.02, 0.04) | (0.01, 0.04) | (96.1, 98.7) |
| $\hat{R}^{S(i)}_{t,Ensemble,i}$ vs. | Median | 0.123 | 0.100 | 90.5 | 0.075 | 0.066 | 93.8 |
| $\hat{R}_{t,Ensemble,i}$ | Range | (0.09, 0.14) | (0.08, 0.11) | (89.6, 92.8) | (0.06, 0.13) | (0.06, 0.12) | (90.1, 94.7) |

Comparison of two-step spatial (covariate-adjusted) INLA prediction, $\hat{R}^{S(i)}_{t,m,i}$, with initial estimates, $\hat{R}_{t,m,i}$, using the leave-one-out validation approach during Wave 2 (between December 16, 2020 – March 02, 2021). The prediction accuracy is presented for counties–Charleston, Greenville, Horry, and Richland and ZIP codes – 29605, 29642, 29680, and 29681. For example, the first row compares the accuracy of the predicted $R_t$ values, $\hat{R}^{S(i)}_{t,Estim,i}$, using proposed two-step spatial (covariate-adjusted) INLA model, where $S(i)$ indicates information from region $i$ is not used in the model fitting in step 2, to their initial estimates, $\hat{R}_{t,Estim,i}$, for these selected counties and ZIP codes.

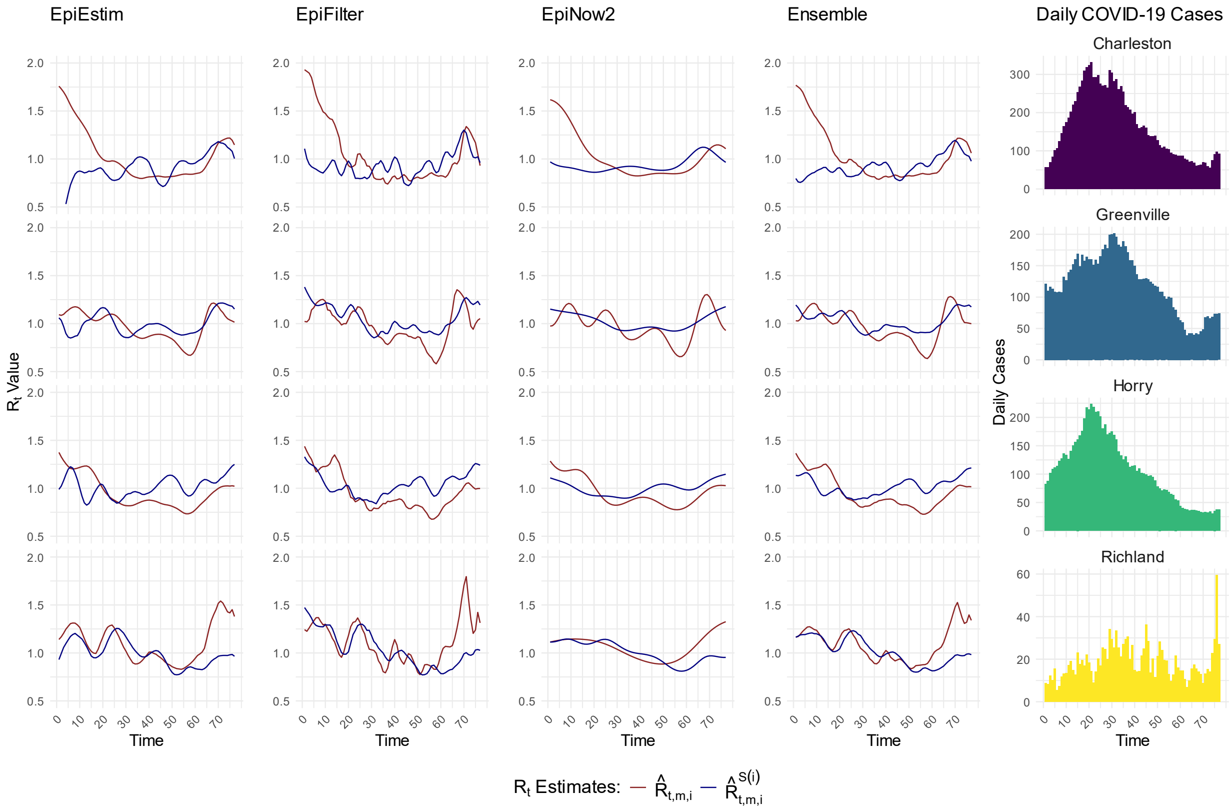


**S4 Fig.** Comparison of two-step spatial (covariate-adjusted) INLA prediction with the initial $R_t$ estimates at the county level in Wave 1.

$\hat{R}^{S(i)}_{t,m,i}$ (blue line) represents the two-step spatial (covariate-adjusted) INLA prediction of effective reproductive number, $\hat{R}^{S(i)}_{t,Estim,i}$, $\hat{R}^{S(i)}_{t,Filter,i}$, $\hat{R}^{S(i)}_{t,Now,i}$ , $\hat{R}^{S(i)}_{t,Ensemble,i}$ while $\hat{R}_{t,m,i}$ (red line) represents the initial estimates $\hat{R}_{t,Estim,i}$, $\hat{R}_{t,Filter,i}$, $\hat{R}_{t,Now,i}$, $\hat{R}_{t,Ensemble,i}$, for select Charleston, Greenville, Horry, and Richland counties during COVID 19 Wave 1 (between June 16, 2020 – August 31, 2020) in SC. Here, $S^{(i)}$ *in* $\hat{R}^{S(i)}_{t,Estim,i}$ indicates that geographic region $i$ was not included in the INLA model fitting. For example, if $i$=*Greenville (GVL),* $\hat{R}^{S(GVL)}_{t,Estim,GVL}$ means Greenville County was not used in the spatial (and covariate-adjusted) INLA model fitting in step 2. The plots in the right most panel present the average daily cases for the respective ZIP codes over the same period.

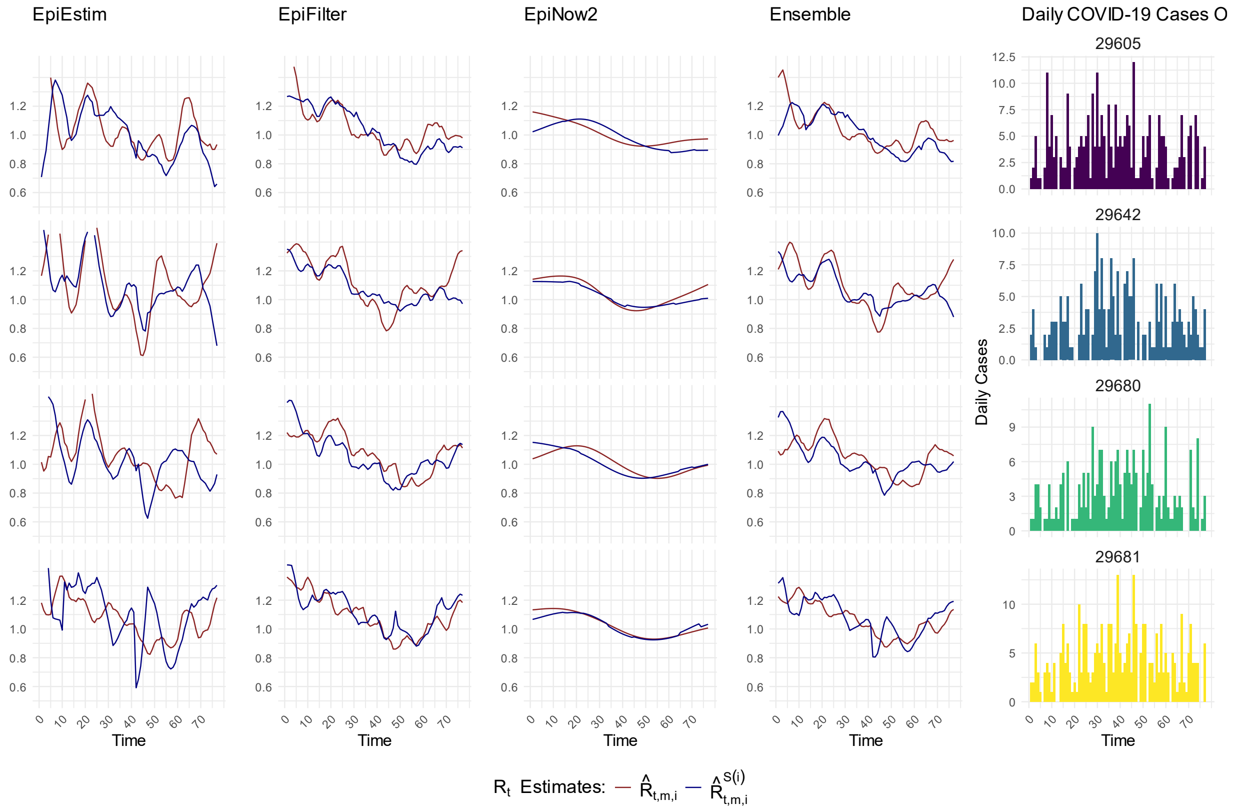


**S5 Fig.** Comparison of two-step spatial (covariate-adjusted) INLA prediction with the initial $R_t$ estimates at the ZIP code level in Wave 1.

$\hat{R}^{S(i)}_{t,m,i}$ (blue line) represents the two-step spatial (covariate-adjusted) INLA prediction of effective reproductive number, $\hat{R}^{S(i)}_{t,Estim,i}$, $\hat{R}^{S(i)}_{t,Filter,i}$, $\hat{R}^{S(i)}_{t,Now,i}$, $\hat{R}^{S(i)}_{t,Ensemble,i}$, while $\hat{R}_{t,m,i}$ (red line) represents the initial estimates $\hat{R}_{t,Estim,i}$, $\hat{R}_{t,Filter,i}$, $\hat{R}_{t,Now,i}$, $\hat{R}_{t,Ensemble,i}$, for select- 29605, 29642, 29680, and 29681 ZIP codes during COVID 19 Wave 1 (between June 16, 2020 – August 31, 2020) in SC. Here, $S(i)$ in $\hat{R}^{S(i)}_{t,Estim,i}$ indicates that geographic region $i$ was not included in the INLA model fitting. For example, if *i=29605,* $\hat{R}^{S(29605)}_{t,Estim,29605}$ means ZIP code "29605" was not used in the spatial (and covariate-adjusted) INLA model fitting in step 2. The plots in the right most panel present the average daily cases for the respective ZIP codes over the same period.

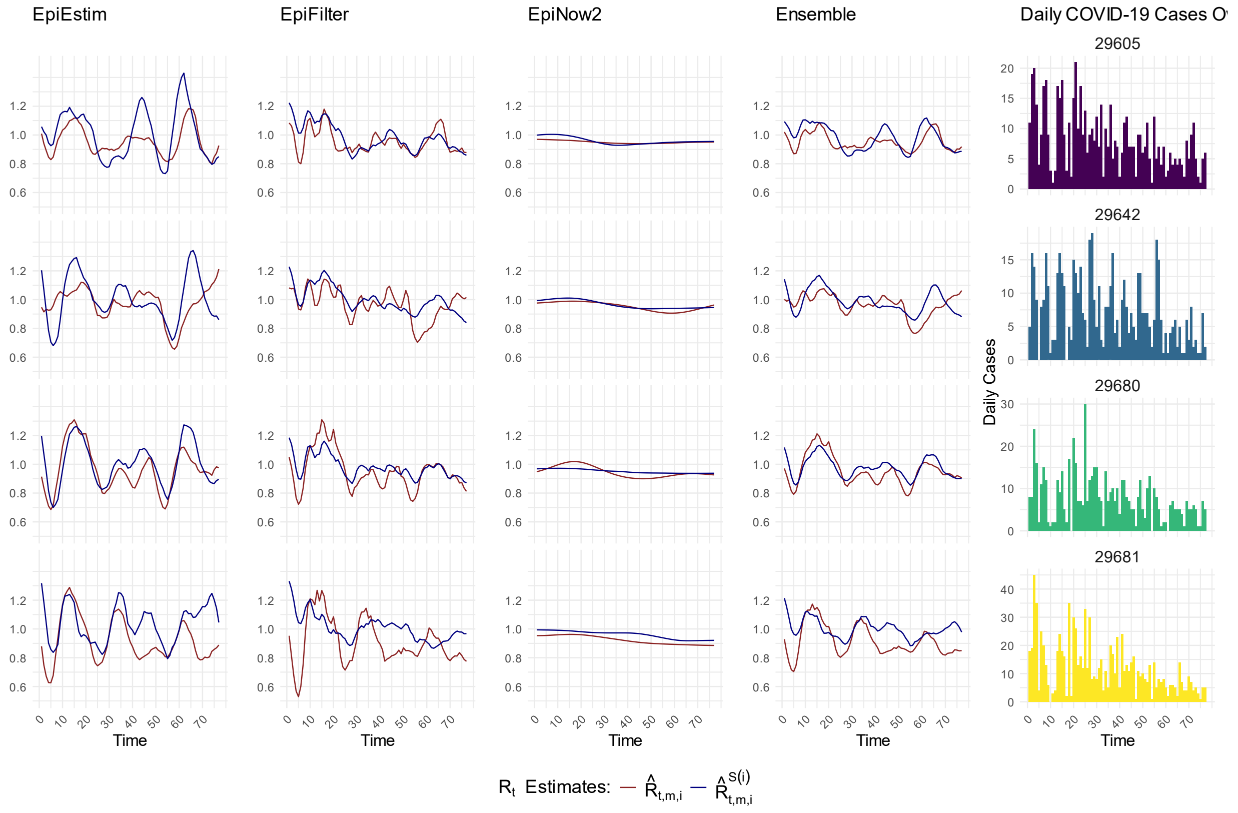


**S6 Fig.** Comparison of two-step spatial (covariate-adjusted) INLA prediction with the initial $R_t$ estimates at the ZIP code level in Wave 2.

$\hat{R}^{S(i)}_{t,m,i}$ (blue line) represents the two-step spatial (covariate-adjusted) INLA prediction of effective reproductive number, $\hat{R}^{S(i)}_{t,Estim,i}$, $\hat{R}^{S(i)}_{t,Filter,i}$, $\hat{R}^{S(i)}_{t,Now,i}$, $\hat{R}^{S(i)}_{t,Ensemble,i}$, while $\hat{R}_{t,m,i}$ (red line) represents the initial estimates $\hat{R}_{t,Estim,i}$, $\hat{R}_{t,Filter,i}$, $\hat{R}_{t,Now,i}$, $\hat{R}_{t,Ensemble,i}$, for select- 29605, 29642, 29680, and 29681 ZIP codes during COVID 19 Wave 2 (between December 16, 2020 – March 02, 2021) in SC. Here, $S(i)$ in $\hat{R}^{S(i)}_{t,Estim,i}$ indicates that geographic region $i$ was not included in the INLA model fitting. For example, if *i=29605,* $\hat{R}^{S(29605)}_{t,Estim,29605}$ means ZIP code "29605" was not used in the spatial (and covariate-adjusted) INLA model fitting in step 2. The plots in the right most panel present the average daily cases for the respective ZIP codes over the same period.

.

**Figures S7, S8:**

**Two-step spatial (covariate-adjusted) INLA estimates ($\hat{R}^S_{t,m,i}$, blue line), $\hat{R}^S_{t,Estim,i}$, $\hat{R}^S_{t,Filter,i}$, $\hat{R}^S_{t,Now,i}$, $\hat{R}^S_{t,Ensemble,i}$ in Wave 2 (between December 16, 2020 – March 02, 2021), and initial estimates ($\hat{R}_{t,m,i}$, red line), $\hat{R}_{t,Estim,i}$, $\hat{R}_{t,Filter,i}$, $\hat{R}_{t,Now,i}$, $\hat{R}_{t,Ensemble,i}$, at the county level in SC. We present the results of the neighboring counties of Greenville and Charleston as examples from the counties used for the second validation approach for prediction.**

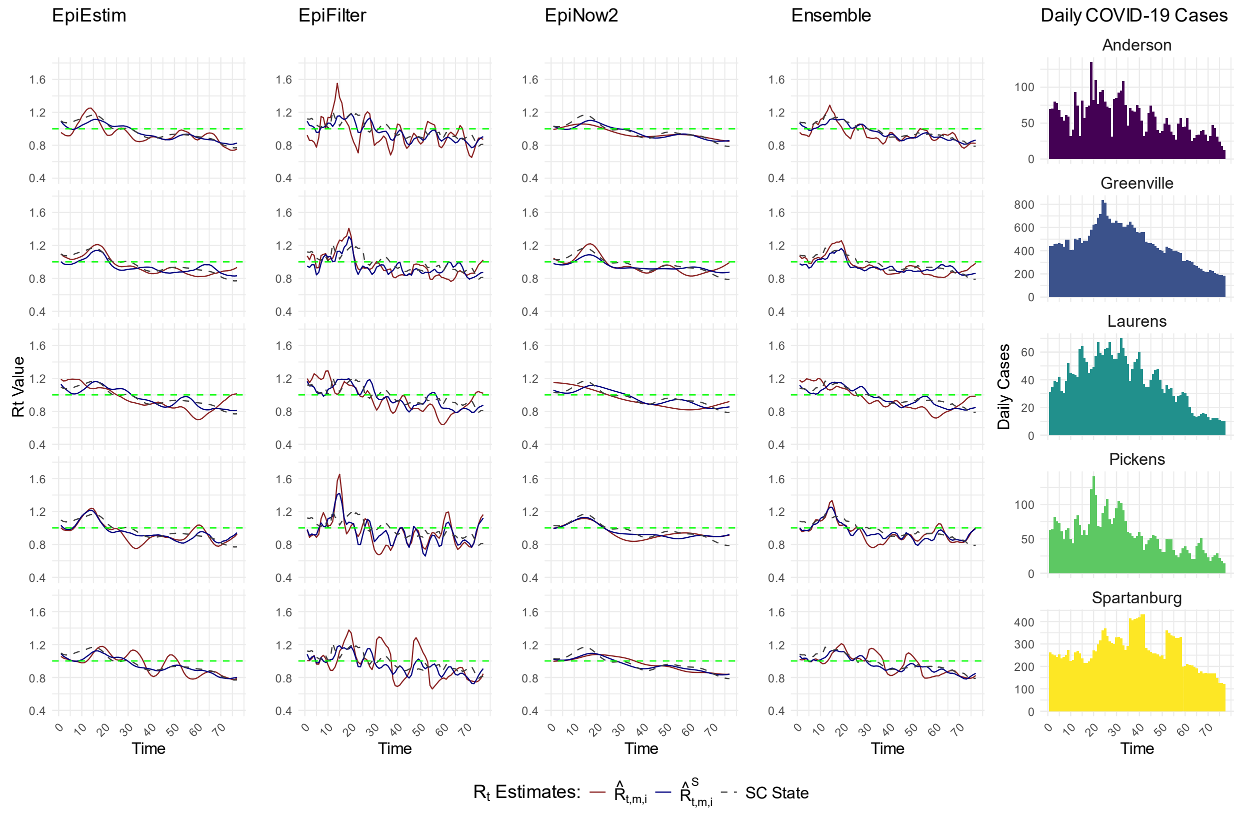


**S7 Fig.** Comparison of $R_t$ estimates of the county Greenville and its neighboring counties in Wave 2. $\hat{R}^S_{t,m,i}$ (blue line) represents the two-step spatial (covariate-adjusted) INLA smoothed estimates, $\hat{R}^S_{t,Estim,i}$, $\hat{R}^S_{t,Filter,i}$, $\hat{R}^S_{t,Now,i}$, and $\hat{R}^S_{t,Ensemble,i}$, while $\hat{R}_{t,m,i}$ (red line) represents the initial estimates, $\hat{R}_{t,Estim,i}$, $\hat{R}_{t,Filter,i}$, $\hat{R}_{t,Now,i}$, and $\hat{R}_{t,Ensemble,i}$, for the county Greenville and its neighboring counties (Anderson, Laurens, Pickens, Spartanburg) during COVID 19 Wave 2 (between December 16, 2020 – March 02, 2021) in SC. The state level initial estimate of $R_t$ is presented with dashed gray line (SC State). The plots in the rightmost panel present the average daily cases for the respective counties over the same period.

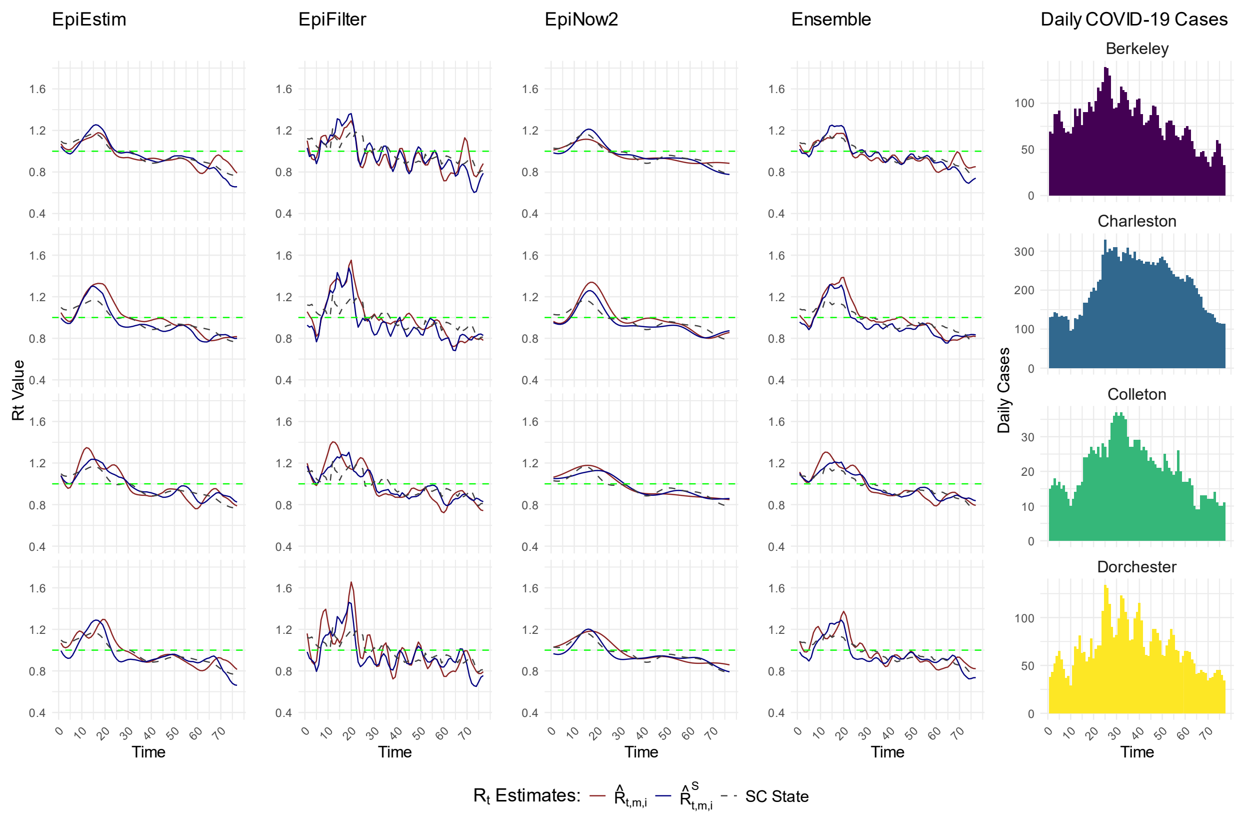


**S8 Fig.** Comparison of $R_t$ estimates of the county Greenville and its neighboring counties in Wave 2.

$\hat{R}^S_{t,m,i}$ (blue line) represents the two-step spatial (covariate-adjusted) INLA smoothed estimates, $\hat{R}^S_{t,Estim,i}$, $\hat{R}^S_{t,Filter,i}$, $\hat{R}^S_{t,Now,i}$and $\hat{R}^S_{t,Now,i}$, while $\hat{R}_{t,m,i}$ (red line) represents the initial estimates, $\hat{R}_{t,Estim,i}$, $\hat{R}_{t,Filter,i}$, $\hat{R}_{t,Now,i}$, and $\hat{R}_{t,Ensemble,i}$, for the county Charleston and its neighboring counties (Berkeley, Colleton, Dorchester) during COVID 19 Wave 2 (between December 16, 2020 – March 02, 2021) in SC. The state level initial estimate of $R_t$ is presented with dashed gray line (SC State). The plots in the rightmost panel present the average daily cases for the respective counties over the same period.

**Table S5:**

**Comparison of $R_t$ estimates between Wave 1 (June 16, 2020 – August 31, 2020) and Wave 2 (December 16, 2020 – March 02, 2021).**

**S5 Table.** Comparison of initial estimates and two-step INLA spatial (covariate-adjusted) estimates between Wave 1 and Wave 2.

| Comparison | County Level (PA (%)) Median (IQR) | | | | ZIP Code Level (PA (%)) Median (IQR) | | | |
|---|---|---|---|---|---|---|---|---|
| | EpiEstim | EpiFilter | EpiNow2 | Ensemble | EpiEstim | EpiFilter | EpiNow2 | Ensemble |
| $\hat{R}_{t,m,i}$ (Wave 1) vs. $\hat{R}_{t,m,i}$ (Wave 2) | 83.79 (81.7, 87.2) | 82.10 (79.9, 84.3) | 88.42 (85.7, 91.0) | 85.60 (82.51, 88.04) | 79.93 (78.0, 81.3) | 86.40 (85.0, 87.1) | 92.29 (91.4, 93.0) | 86.71 (85.5, 87.6) |
| $\hat{R}^S_{t,m,i}$ (Wave 1) vs. $\hat{R}^S_{t,m,i}$ (Wave 2) | 87.43 (86.0, 89.0) | 86.62 (85.4, 87.7) | 90.96 (90.3, 91.9) | 88.53 (87.34, 89.75) | 85.62 (83.5, 87.4) | 90.35 (89.3, 91.3) | 93.78 (93.0, 95.0) | 90.31 (89.8, 91.4) |

Comparison of initial estimates, $\hat{R}_{t,m,i}$ and two-step spatial INLA (covariate-adjusted) estimates ,$\hat{R}^S_{t,m,i}$, between COVID-19 Wave 1 (between June 16, 2020 – August 31, 2020) and Wave 2 (between December 16, 2020 – March 02, 2021) at the county and ZIP code levels in SC for each method m (i.e. EpiEstim, EpiFilter, EpiNow2, and Ensemble).

#### Simulation Study: Generation of $R_t$ and COVID-19 Case Counts at the County Level in South Carolina (SC)

##### Effective Reproductive Number ($R_t$) Generation

In this simulation study, we generate the effective reproductive number ($R_t$) and then use it for COVID-19 daily case counts generation for the counties in SC. The $R_t$ generation process incorporates spatial correlations and epidemiological considerations. The time-varying effective reproductive number, $R_{t,i}$, for each county $i$ at time $t$, is generated using a sinusoidal function to reflect the periodic variations in disease transmissions:

$$R_{t,i} = \beta_{0,i} + \beta_{1,i} \sin(t.\beta_{2,i}) + \beta_{3,i}$$

where, $\beta_{0,i}$ is baseline reproductive number, representing the average $R_t$ value for a county without temporal variation, amplitude of the sinusoidal component is $\beta_{1,i}$, frequency of the sinusoidal component is $\beta_{2,i}$, and the term $\beta_{3,i}$ is linear adjustment.

Each parameter $\boldsymbol{\beta_0}, \boldsymbol{\beta_1}, \boldsymbol{\beta_2}, \boldsymbol{\beta_3}$ is generated separately for $N$ counties from a multivariate normal (MVN) distribution:

$$\boldsymbol{\beta_k} = (\beta_{k,1}, \beta_{k,2}, \dots, \beta_{k,N})^T \in \boldsymbol{MVN}(\boldsymbol{0}, \boldsymbol{\Sigma}), \quad k \in \{0,1,2,3\}$$

where, $\boldsymbol{\beta_k}$ represents a vector of size $N$, corresponding to a specific parameter for all counties. The covariance matrix $\boldsymbol{\Sigma} \in \mathbb{R}^{N \times N}$ captures the spatial correlation among the counties. We define $\boldsymbol{\Sigma} = (\boldsymbol{D} + 0.8 * \boldsymbol{W})$, where $\boldsymbol{D}$ is an identity matrix, and $\boldsymbol{W}$ is the adjacency matrix, where $w_{i,j} = 1$ if counties $i, j$ share a border and $w_{i,j} = 0$ otherwise.

Since the values of $\boldsymbol{\beta_k}$ are drawn from the MVN distribution may not fall within meaningful epidemiological ranges, we apply a scaling transformation:

$$\boldsymbol{\beta_k^*} = \frac{\boldsymbol{\beta_k} - a_k}{c_k} \times (u_k - l_k) + l_k = \frac{u_k - l_k}{c_k}\boldsymbol{\beta_k} - \frac{a_k}{c_k} \times (u_k - l_k) + l_k\,, \quad k \in \{0,1,2,3\}$$

where $a_k = \min(\boldsymbol{\beta_k})\,, c_k = \max(\boldsymbol{\beta_k}) - \min\,(\boldsymbol{\beta_k})$, $l_k$ and $u_k$ represent the lower and upper bounds for the corresponding parameter. The transformation ensures that the parameters $\boldsymbol{\beta_k^*}$ fall within predefined epidemiological bounds: $\boldsymbol{\beta}_{\boldsymbol{k}}^{*} \in [l_k, u_k]$. Thus, after transformation, the final $4 \times N$ matrix of parameters follows:

$$\boldsymbol{\beta}_{\boldsymbol{k}}^{*} = \left(\beta_{k,1}^{*}, \beta_{k,2}^{*}, \dots, \beta_{k,N}^{*}\right)^{T} \sim \boldsymbol{MVN}\left(\left(-\frac{a_k}{c_k} \times (u_k - l_k) + l_k\right)\mathbf{1}, \frac{u_k - l_k}{c_k}\boldsymbol{\Sigma}\right), \quad k \in \{0, 1, 2, 3\}$$

The baseline reproductive number, $\boldsymbol{\beta}_{\mathbf{0}}^{*}$, is constrained between 0.9 and 1.2. The amplitude of the sinusoidal component, $\boldsymbol{\beta}_{\mathbf{1}}^{*}$, ranges from 0.2 to 0.3, controlling the magnitude of the waves of the disease. The frequency of the sinusoidal component, $\boldsymbol{\beta}_{\mathbf{2}}^{*}$, ranges between 0.3 and 0.5, and is further adjusted using $2\pi/(45 + \boldsymbol{\beta}_{\mathbf{2}}^{*} \times 50)$ to determine wave periodicity. The linear adjustment term, $\boldsymbol{\beta}_{\mathbf{3}}^{*}$, ranges from -0.2 to 0.1, accounting for the additional shifts in $R_t$. We constrain $R_t$ values between 0.8 and 1.5 to avoid large fluctuations.

Using these transformed parameters, the final $R_t$ values for each county $i$ at each time point $t$ are computed as:

$$R_{t,i} = \beta_{0,i}^{*} + \beta_{1,i}^{*} \sin\left(t.\beta_{2,i}^{*}\right) + \beta_{3,i}^{*}$$

**COVID-19 Case Generation**

To simulate daily COVID-19 case counts at the county level in SC, we incorporate the serial interval distribution and the effective reproductive number $R_{t,i}$. The serial interval, representing the time between the symptom onset of a primary case and a secondary case, was assumed to follow a gamma distribution. We use a discretized gamma distribution with a mean of 4.7 days and a standard deviation of 2.9 days. We initialize each county with 200 past cases, which are evenly distributed over the past 10 days. The past cases and normalized serial interval distribution are used to compute the total infectiousness ($\Lambda_{t,i}$) at each time $t$ and location $i$, given by:

$$\Lambda_{t,i} = \sum_{k=1}^{s} I_{t-k,i} p_k,$$

where, $I_{t-k,i}$ is the number of past cases on day $t - k$, $p_k$ represents the probability that a primary case generates a secondary case between $k - 1$ and $k$ days, based on the discretized gamma distribution, $s$ is the maximum infectious period considered in the model. Using the generated effective reproductive number $R_{t,i}$ for day $t$, the expected number of new cases at location $i$, ($\mathrm{E}[I_{t,i}]$), is given by:

$$\mathrm{E}\left[I_{t,i}\right] = \Lambda_{t,i} \times R_{t,i},$$

Daily case counts are drawn from a Poisson distribution:

$$I_{t,i} \sim Poisson\left(\Lambda_{t,i} \times R_{t,i}\right).$$

We generate 50 independent sets of daily COVID-19 case data at the county level in SC with the same $R_{t,i}$. For each simulation, we generate 140 time points and perform initial estimation of $R_{t,i}$ using EpiEstim, EpiFilter, and an ensemble-based estimation approach, while excluding EpiNow2 due to its high computational cost. The analysis workflow closely flows the approach used for real data, including initial $R_t$ estimation, two-step spatial (covariate-adjusted) INLA smoothing of $R_t$, and prediction of $R_t$ for regions with entirely missing data.

We perform initial estimation over the full time series but discard the first 40 data points as burn-in period to ensure that the analysis is not influenced by initialization effects. To make the prediction of $R_t$ for the regions with entirely missing data, we systematically leave a selected county out from the INLA model fitting process, making its data unavailable. The INLA model is then trained using data from the remaining counties, and $R_t$ is predicted for the excluded county. This process is repeated for multiple counties to assess the reliability of $R_t$ estimation for the counties with entirely missing data. Unlike real data, where the true $R_t$ is unknown, the simulation study provides generated $R_t$ values, allowing us to direct benchmark and compare the performance of different estimation methods.

### Simulation Results

We evaluated the performance of our two-step spatial (covariate-adjusted) INLA smoothing and prediction framework for estimating the effective reproductive number ($R_t$) at the county level. The estimates were compared with those obtained from existing methods, including EpiEstim, EpiFilter, and an ensemble-based estimation approach. The analysis was conducted using simulated COVID-19 case data for counties in SC and focused on evaluating the accuracy of initial estimates, the improvements achieved through spatial smoothing, and the predictive ability of the framework for the counties with completely missing data.

Figure **S9** presents a comparison between the initial $R_t$ estimates obtained using EpiEstim, EpiFilter, and the ensemble approach, the smoothed $R_t$ estimates generated through the two-step spatial (covariate-adjusted) INLA framework, and the true $R_t$ values used in the simulation. The results demonstrate that the initial estimates are close to the generated $R_t$ across counties. However, after applying the spatial INLA smoothing technique, the estimates align more closely with the true $R_t$ values. The percentage agreement (PA) values in Table **S6** further confirm that the smoothed $R_t$ estimates provide a highly accurate representation of the true values. In this scenario, all available data were used, meaning that the initial estimates were available for all counties before the spatial INLA model was applied.

Figure **S10** evaluates the predictive capability of our approach for estimating $R_t$ in counties with completely missing data. The estimation process involved systematically excluding each selected county one by one from model fitting. For example, Charleston County was left out to simulate a scenario where data for that county was unavailable. The model then predicted $R_t$ for the missing county by borrowing information from neighboring counties. This procedure was repeated for Greenville, Horry, and Richland counties. The results indicate that the predicted $R_t$ values from the INLA model show strong agreement with the true generated $R_t$ values, demonstrating the robustness of the method. The percentage agreement values in Table **S6** further validate that the proposed framework maintains high predictive performance, even for counties with entirely missing data.

**Figures S9, S10, and Table S6:**

**Comparison of the $R_t$ estimates using the existing methods, including EpiEstim, EpiFilter and an Ensemble based approach, with our two-step spatial (covariate-adjusted) INLA smoothing and prediction of $R_t$ for areas with completely missing data, benchmarked against the true $R_t$ values generated for counties in South Carolina (SC).**

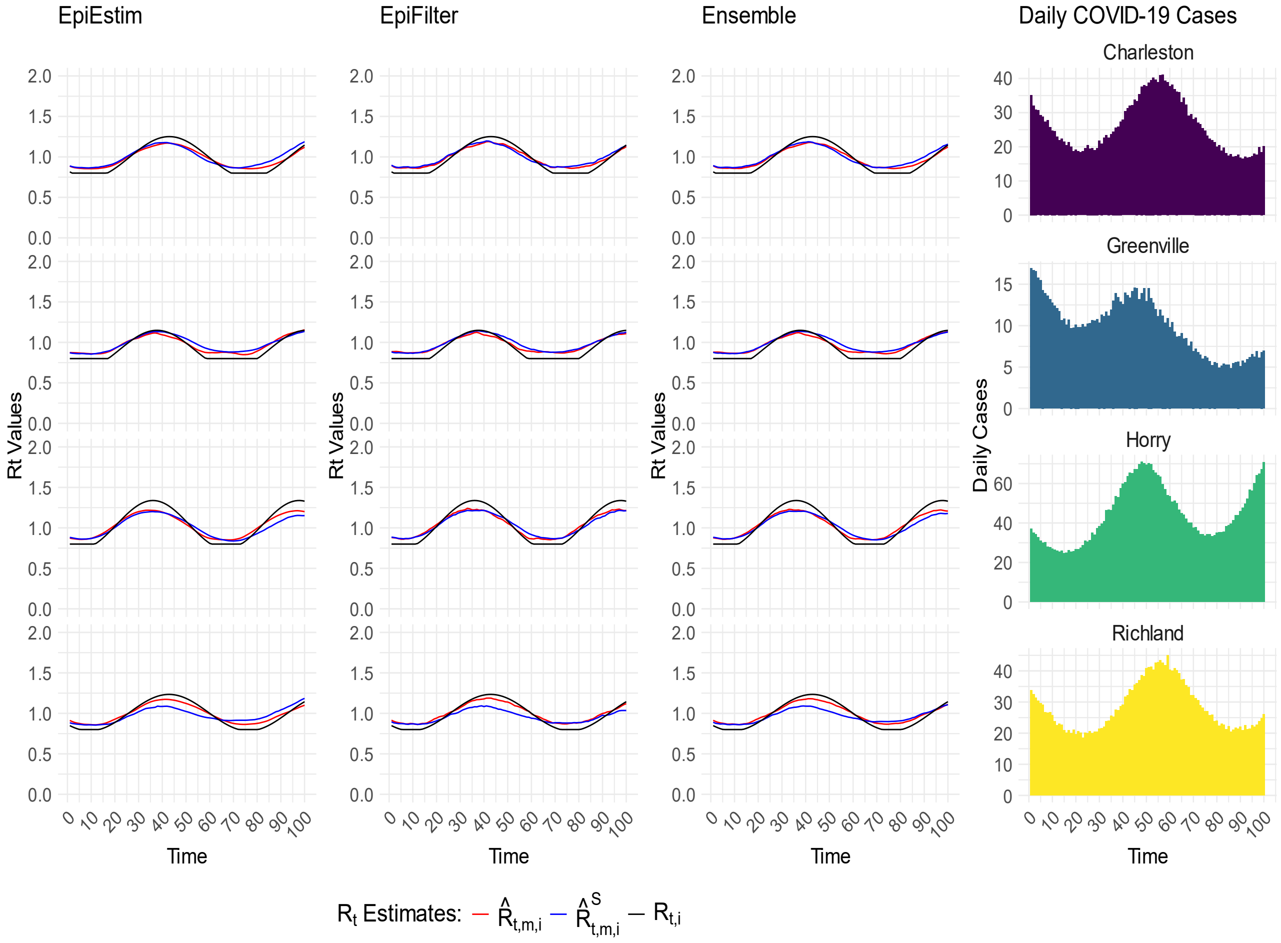


**S9 Fig.** Comparison of two-step spatial (covariate-adjusted) INLA smoothing and initial $R_t$ estimates with the generated true $R_t$.

$\hat{R}^S_{t,m,i}$ (blue line) represents the two-step spatial (covariate-adjusted) INLA smoothed estimates, $\hat{R}^S_{t,Estim,i}$, $\hat{R}^S_{t,Filter,i}$, and $\hat{R}^S_{t,Ensemble,i}$. Similarly, $\hat{R}_{t,m,i}$ (red line) denotes the initial estimates, $\hat{R}_{t,Estim,i}$, $\hat{R}_{t,Filter,i}$, and $\hat{R}_{t,Ensemble,i}$, while $R_{t,i}$ represents the true generated $R_t$ at location $i$. These estimates are presented for selected counties in SC (Charleston, Greenville, Horry, and Richland) based on simulated COVID-19 case data. The rightmost panel illustrates the average daily cases were calculated from the 50 simulated datasets.

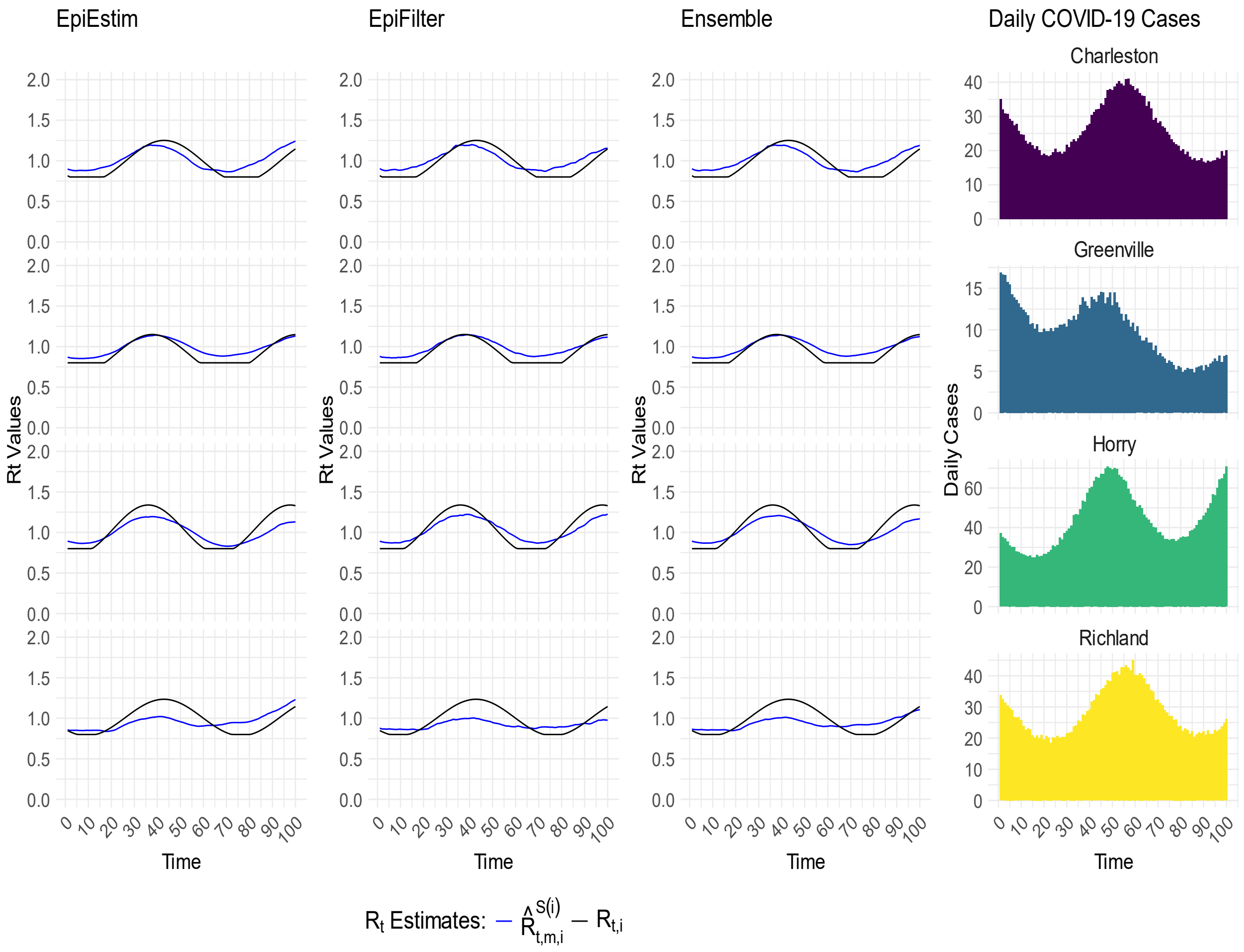


**S10 Fig.** Comparison of two-step spatial (covariate-adjusted) INLA prediction with the generated true $R_t$. $\hat{R}^{S(i)}_{t,m,i}$ (blue line) represents the two-step spatial (covariate-adjusted) INLA prediction of effective reproductive number, $\hat{R}^{S(i)}_{t,Estim,i}$, $\hat{R}^{S(i)}_{t,Filter,i}$, $\hat{R}^{S(i)}_{t,Ensemble,i}$, while $R_{t,i}$ (black line) represents the generated true $R_t$ values for selected counties (Charleston, Greenville, Horry, and Richland) in SC. In this setting, we assumed the data for the selected counties were completely missing, making it impossible to estimate $R_t$ using the existing methods. However, our two-step spatial (covariate-adjusted) INLA framework borrowed information from neighboring counties to predict $R_t$ for these areas. Here, $S(i)$ in $\hat{R}^{S(i)}_{t,Estim,i}$ indicates that geographic region $i$ was not included in the INLA model fitting. For example, if $i$=*Greenville (GVL)*, $\hat{R}^{S(GVL)}_{t,Estim,GVL}$ means Greenville County was not used in the spatial (covariate-adjusted) INLA model fitting in step 2. The rightmost panel illustrates the average daily cases were calculated from the 50 simulated datasets.

**S6 Table.** Comparison of initial estimates, two-step spatial (covariate-adjusted) INLA smoothed estimates, and predicted $R_t$ with the true generated $R_t$.

| Comparison | Accuracy Measurement Metrics Median (IQR) | | |
|---|---|---|---|
| | RMSE | MAPE | PA (%) |
| $\hat{R}_{t,Estim,i}$ vs. $R_{t,i}$ | 0.118 (0.114-0.122) | 0.085 (0.082-0.088) | 92.55 (92.63-93.74) |
| $\hat{R}^{S}_{t,Estim,i}$ vs. $R_{t,i}$ | 0.081 (0.072-0.091) | 0.076 (0.066-0.084) | 92.96 (92.23-92.73) |
| $\hat{R}^{S(i)}_{t,Estim,i}$ vs. $R_{t,i}$ | 0.118 (0.109-0.145) | 0.100 (0.092-0.112) | 90.76 (89.67-91.38) |
| $\hat{R}_{t,Filter,i}$ vs. $R_{t,i}$ | 0.195 (0.195-0.206) | 0.106 (0.101-0.110) | 91.52 (91.12-91.83) |
| $\hat{R}^{S}_{t,Filter,i}$ vs. $R_{t,i}$ | 0.081 (0.074-0.088) | 0.075 (0.067-0.083) | 93.10 (92.43-93.75) |
| $\hat{R}^{S(i)}_{t,Filter,i}$ vs. $R_{t,i}$ | 0.110 (0.103-0.115) | 0.094 (0.090-0.100) | 91.30 (90.79-91.57) |
| $\hat{R}_{t,Ensemble,i}$ vs. $R_{t,i}$ | 0.154 (0.151-0.160) | 0.094 (0.090-0.098) | 92.09 (91.72-92.37) |
| $\hat{R}^{S}_{t,Ensemble,i}$ vs. $R_{t,i}$ | 0.081 (0.073-0.088) | 0.075 (0.065-0.083) | 93.10 (92.47-93.91) |
| $\hat{R}^{S(i)}_{t,Ensemble,i}$ vs. $R_{t,i}$ | 0.113 (0.108-0.122) | 0.096 (0.092-0.107) | 91.14 (90.33-91.40) |

The table presents a comparison of accuracy measurement metrics for $R_t$ estimates obtained using various methods against the true generated $R_t$ values. Metrics include the root mean squared error (RMSE), mean absolute prediction error (MAPE), and percentage agreement (PA), along with their median and interquartile range (IQR). The initial estimates, $\hat{R}_{t,Estim,i}$, $\hat{R}_{t,Filter,i}$, and $\hat{R}_{t,Ensemble,i}$, were obtained using the existing techniques and an ensemble based estimation. $\hat{R}^{S}_{t,Estim,i}$, $\hat{R}^{S}_{t,Filter,i}$, and $\hat{R}^{S}_{t,Ensemble,i}$ represent the two-step spatial (covariate-adjusted) INLA estimates. The predicted estimates ($\hat{R}^{S(i)}_{t,Estim,i}$, $\hat{R}^{S(i)}_{t,Filter,i}$, $\hat{R}^{S(i)}_{t,Ensemble,i}$) represent $R_t$ values for areas with completely missing data. In these cases, data for the selected counties (Charleston, Greenville, Horry, and Richland) were assumed to be unavailable, making it impossible to estimate $R_t$ using the existing methods. However, our two-step spatial (covariate-adjusted) INLA framework borrowed information from neighboring counties to predict $R_t$ for these areas. These estimates were benchmarked against the generated true $R_t$ values.
